# BenchmarkHub: an open-source platform for reproducible evaluation of large language models in healthcare

**DOI:** 10.64898/2026.11.11.26367866

**Authors:** Juan Ignacio Alvarez-Arenas, Daniel Jimenez-Carretero, Fátima Sánchez-Cabo

## Abstract

**Background:** Large language models (LLMs) are solely judged by their scores on biomedical benchmarks, yet these scores are rarely reproducible since the prompts, inference settings and model builds are seldom reported. We developed BenchmarkHub, an open-source platform for auditable evaluation of language and vision-language models for biomedical research.

*Methods:* BenchmarkHub is a Python web application (Django 5.2, SQLite) released under the MIT licence and available at https://github.com/juanjuanignacio/benchmarkhub. The application can be accessed through a browser interface, a set of command-line tools and a REST API, so interactive exploration and scripted pipelines produce identical, comparable records. The platform samples questions with fixed seeds, runs models directly or as tool-using agents in a code sandbox, grades answers with deterministic rules and stores every prompt, reply and tool call. The interface provides a leaderboard to rank performance of the models, allowing model comparison without any preexisting programming knowledge.

*Findings:* We used BenchmarkHub to evaluate 12 open-weight models in five experiments on 15 biomedical benchmarks (11 text, two multimodal and two computational biology) and two general benchmarks. The pooled accuracy ranged from 46·1% (95% CI 43·0–49·3) to 80·9% (78·3–83·3). Re-grading the responses with simple answer-extraction rules lowered a model’s accuracy by up to 33·1 percentage points leading to a reorder of the leaderboard. Published scores were found for 23 out of the 143 benchmark/model combinations tested. The accuracy measured by BenchmarkHub was lower than in the published results in 19 out of the 23 available comparisons. The effect of four prompting protocols (1: default prompt; 2: asking the model to answer the letter only; 3: extended reasoning; 4: three-shot prompting) changed MEDQA accuracy by up to 11·3 points but was not significant after multiple-testing correction with 150 questions. We also tested the impact of ablation on benchmarking using the MMMU benchmark dataset. Removing images lowered the accuracy by 18·0 and 19·3 points repectively for the two strongest models. Finally, the accuracy of agentic benchmarks using a sandboxed interpreter increased on computational biology tasks by 36–63 points.

*Interpretation:* Benchmarking is essential to assess the impact and validity of LLMs for medical applications. BenchmarkHub allows a fair and rigorous comparison of models across established datasets, including the possibility to upload own datasets. The suite includes visual tools to ease model comparison and ranking without any programming knowledge. Self-hosted infrastructure that records every choice and parameter makes benchmark claims checkable without data leaving the institution.

*Funding:* European Union EU4Health Programme (JACARDI), Spanish Ministry of Science, Innovation and Universities–State Research Agency (MCIN/AEI) with FEDER funds, the European Union Recovery and Resilience Facility–NextGenerationEU, and the Instituto de Salud Carlos III.

*Research in context:* 

*Evidence before this study:* We searched PubMed, arXiv and Crossref in October, 2026, for articles published from January, 2022, to October, 2026, without language restrictions, using combinations of the terms “large language model”, “benchmark”, “evaluation”, “reproducibility”, “prompt sensitivity”, “medical”, “clinical”, “multimodal”, “image”, “agent” and “bioinformatics”. A systematic review of 519 studies found that LLM evaluations in healthcare rely mainly on exam-style questions, rarely use real patient data and seldom report evaluation settings. Holistic frameworks and new task-specific benchmarks, including two published in this journal have broadened what is measured, but headline scores are usually produced by model developers with incompletely described protocols. Studies outside medicine show that prompt format, answer extraction and nominally deterministic decoding can each change benchmark accuracy substantially, and that many questions in general multimodal benchmarks can be answered from the text alone, without looking at the image. Much of this evidence comes from preprints or developer-run evaluations, and we found no medical study that quantified, on the same items and within one framework, the effects of prompting protocol, answer extraction, image availability and tool access, nor an open, self-hosted platform built for that purpose.

*Added value of this study:* BenchmarkHub is, to our knowledge, the first open-source platform that combines a web interface, command-line and REST access, local and cloud model providers, a sandboxed tool-use agent loop, procedurally generated computational biology benchmarks and a complete per-question audit trail. Using only the platform and 12 locally served open-weight models, we show that the answer-extraction rule alone can change medical accuracy by more than the gaps between models and reorder them, that published scores are usually higher than those measured under a documented protocol, that protocol effects need larger samples than are typically used, that on a medical multimodal benchmark the strongest models kept about 90% of their accuracy without the images, and that tool access transforms performance on computational biology tasks.

*Implications of all the available evidence:* Medical benchmark scores should be reported together with the protocol, answer-extraction rules, decoding settings, model build and input modality that produced them, and comparisons between models should be paired and made under identical conditions. Self-hosted evaluation infrastructure allows hospitals and research institutes to do this for their own candidate models without sharing data, as a reproducible screening step that precedes, and does not replace, clinical validation.

## Introduction

Large language models (LLMs) currently need to pass medical licensing-style examinations.^1^ Multiple-choice benchmarks such as MedQA, MedMCQA, PubMedQA and the medical subsets of MMLU have become the default evidence offered for their medical competence.^2–5^ A systematic review of 519 evaluation studies found that most assessed accuracy is on exam-style questions and only 5% used real patient data.^6^ Benchmark accuracy also transfers poorly to real-world use: although LLMs identified at least one relevant medical condition in 94·9% of scenarios when assessed independently, members of the public using the same models identified relevant conditions in fewer than 34·5% of cases, performing no better than participants who used other sources of information.^7^ Diagnostic performance also degrades when models must interpret real patient information revealed sequentially.^8^ Calls to retire exam benchmarks as headline evidence^9^ and to validate generative models rigorously before clinical use^10^ have been answered with broader frameworks, such as MedHELM,^11^ and with new benchmarks, including RareArena and CARDBiomedBench, published in this journal.^12,13^

A less discussed problem is that a benchmark score depends not only on the model but on the whole evaluation pipeline: which questions were sampled, how the prompt was worded, how many tokens the model was allowed, how the answer was extracted from free text, which model build was served, and whether additional images or tools were available. Outside medicine, equivalent prompt formats,^14,15^ answer-extraction rules^16^ and nominally deterministic decoding^17^ each change accuracy substantially, and harness developers list these as the main obstacles to reproducibility.^18^ Multimodal benchmarks can often be answered from the text alone,^19^ and models that reach expert-level accuracy on medical image challenges often reason wrongly about the image.^20^ These choices are rarely reported in medical evaluations, and developer-reported scores come from in-house protocols that cannot be rerun.^21^

Existing frameworks address parts of the problem. HELM and the *Language model evaluation harness* provide standardised benchmarks,^18,22^ but they are code-first tools for machine learning engineers, and medical suites are typically run centrally by their developers.^11^ Open-weight models are increasingly competitive on difficult diagnostic cases,^23^ and running them locally keeps sensitive material in the institution, but it makes evaluation the institution’s responsibility.

We developed BenchmarkHub, an open-source, self-hostable platform that makes every step of the evaluation pipeline an explicit, recorded and re-runnable variable, and that extends evaluation from single-turn questions to tool-using agents. Here we describe the platform and report five use cases, run entirely through its public API on locally served open-weight models, that quantify how answer extraction, prompting protocol, image availability and tool access change medical and biomedical results. BenchmarkHub is available at https://github.com/juanjuanignacio/benchmarkhub.

## Methods

### Platform design

BenchmarkHub is a Python web application (Django 5.2, SQLite) released under the MIT licence. The application can be accessed through a browser interface, a set of command-line tools and a REST API, so interactive exploration and scripted pipelines produce identical, comparable records (Figure 1). Models are reached through provider adapters for local inference servers (Ollama, vLLM and any OpenAI-compatible endpoint) and for cloud APIs (OpenAI, Anthropic, Google, Mistral, Groq and Cohere). When a local server is used, prompts, questions and outputs never leave the institution. Per-provider concurrency limits and retry policies protect shared inference servers.

**Figure 1:**
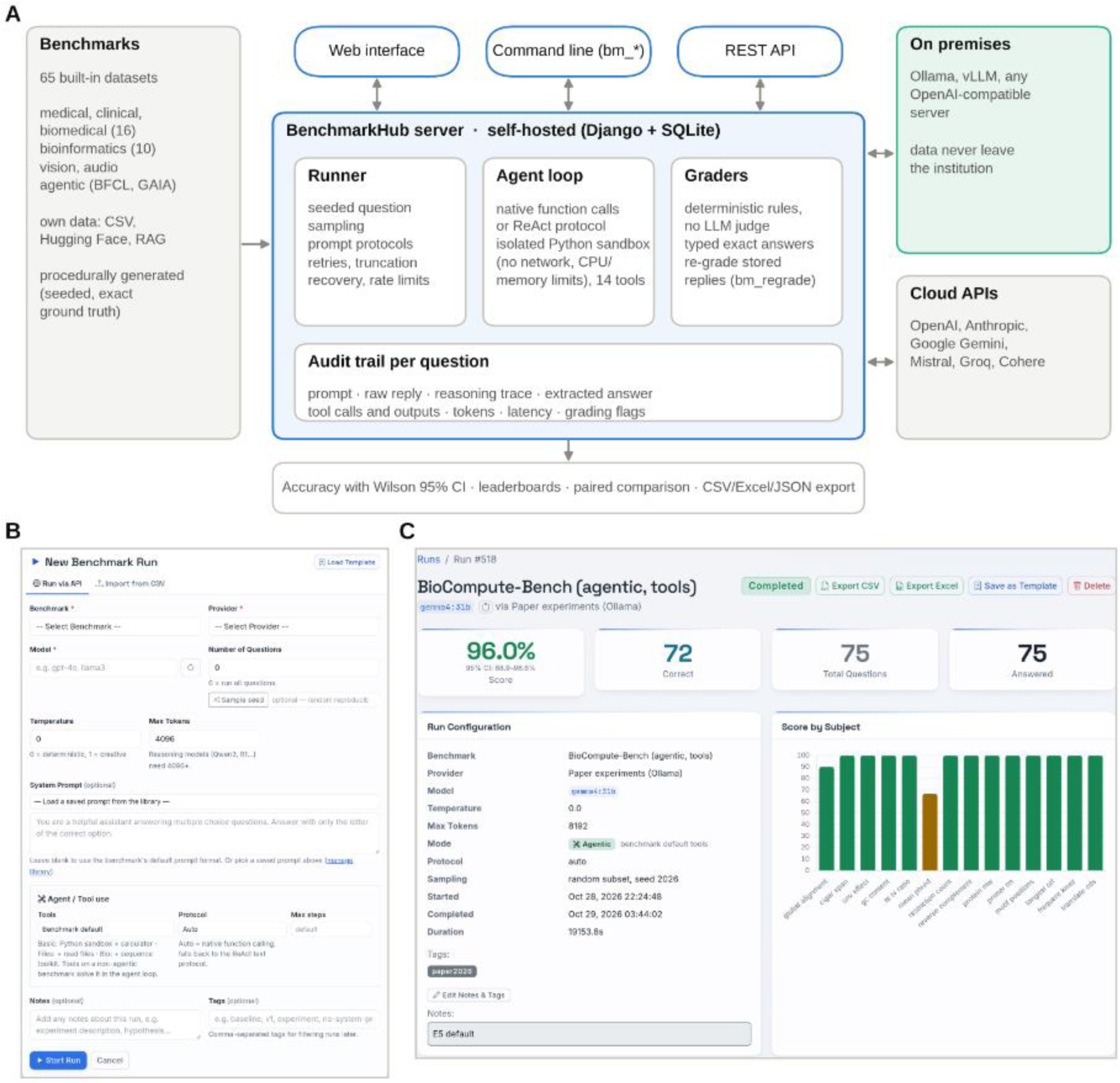
BenchmarkHub architecture and interface. (A) Three interfaces (web, command line and REST API) operate one self-hosted server that samples questions reproducibly, runs single completions or a tool-using agent loop in an isolated sandbox, grades answers with deterministic rules and stores a complete audit trail per question. Models are served on premises (Ollama, vLLM or any OpenAI-compatible server) or through cloud APIs. (B) Run configuration in the web interface, including sample seed, prompting protocol and tool access. (C) A completed run (Gemma 4 31B on BioCompute-Bench with tools) showing accuracy with its Wilson 95% CI, the recorded configuration and the per-family breakdown.

The platform ships loaders for 65 benchmarks, including 16 medical, clinical or biomedical datasets and ten additional ones for bioinformatics questions. Users can add their own questions from CSV files, the Hugging Face Hub or document collections. A run is defined by a benchmark, a model, a sample size with an optional seed (so different models receive the same questions), sampling parameters, a prompting protocol (system instruction, chain-of-thought cue or reproducibly drawn k-shot examples) and, optionally, tools. Vision benchmarks can be run with images removed, as a text-only ablation on identical items.

Questions are answered by a single completion or inside an agent loop that uses the provider’s native function calling or, failing that, a text protocol. Tools (a Python interpreter, a calculator, file reading and eight bioinformatics sequence utilities) run in a sandbox: a separate Python process with CPU-time, memory and file-size limits, no network access and a fresh working directory per question (appendix p 5-6). Every assistant turn, tool call and tool output is stored as an inspectable trajectory.

Answers are graded by deterministic rules; no LLM is used as a judge. Multiple-choice answers are extracted by an ordered set of rules that recognises explicit answer statements and conclusion lines before falling back to isolated option letters, and the procedurally generated benchmarks store a typed answer (a number, sequence, list or label) that is compared exactly or, for decimal values, within a stated tolerance. Reasoning traces are removed before grading; a reply truncated without an answer is retried once, and remaining empty replies are graded on the end of the reasoning trace, with both events flagged. For each question the platform stores the prompt, raw reply, reasoning trace, extracted answer, correctness, tokens and latency, so grading rules can be improved and all runs re-scored without new inference (appendix pp 3–5).

To evaluate computational biology tasks rather than recall, the platform includes two procedurally generated benchmarks: BioCompute-Bench (300 single-step computations in 15 families, eg, translation, open reading frames, variant effect, CIGAR span, alignment score) and BioAgent-Bench (80 multi-step analyses over generated FASTA, VCF, GFF3, FASTQ and expression files, eg, splicing and translating a multi-exon gene). Each item comes from its own seeded generator, so every installation produces identical items that do not exist on the web; ground truth is computed by the code that implements the sequence-analysis tools, and we recomputed all 380 answers independently with Biopython (appendix p 8).

### Models and inference

We evaluated 12 open-weight models served by Ollama (version 0.32.14) on a shared institutional multi-GPU server: seven instruction models (Llama 3.1 8B, Mistral 7B, Qwen2.5 7B, Phi-4 14B, Gemma 3 12B, Gemma 3 27B and Llama 3.3 70B), four models that produce a reasoning trace before answering (DeepSeek-R1 7B, GLM-4.7-Flash, Gemma 4 31B and Qwen3.8 27B) and one additional vision-language model (Qwen2.5-VL 7B). Together they cover the main open model families, sizes from 7 to 70 billion parameters, instruction and reasoning models, and models with published scores on the benchmarks used. Each build is identified by its Ollama digest. All were 4-bit quantised builds (Q4_K_M) except Llama 3.1 8B (16-bit). The context window was 32 768 tokens, temperature 0 unless stated, and the reply limit 2048 tokens for instruction models and 8192 for reasoning models and agent runs (appendix p 1).

### Experiments

All experiments were defined in a declarative plan and submitted as ordinary runs through the REST API by a client script, so every result is visible and auditable in the web interface. Questions were sampled with seed 2026, so every model answered the same items.

The experiments addressed three questions: how locally measured accuracy compares with published scores (experiment 1); how much it varies with evaluation choices, namely answer extraction, prompting protocol and sampling temperature (answer-extraction analysis and experiments 2 and 3); and what models gain from images and tools, assessed by removing them (experiments 4 and 5).

*Experiment 1: biomedical leaderboard and agreement with published scores.* The 11 text models answered 150 biomedical questions each from MedQA (USMLE, four options), MedMCQA, PubMedQA (with the full abstract) and MedXpertQA Text (ten options), and 50 from each of seven medical MMLU subsets (clinical knowledge, medical genetics, anatomy, professional medicine, college biology, college medicine and virology), giving 950 medical questions per model, plus 150 general questions from each of MMLU and MMLU-Pro for comparison purposes.^2–5,24,25^ Prompts were zero-shot platform defaults. We collected every score for these model–benchmark pairs that we could verify in a developer technical report, model card or benchmark paper, together with its evaluation setting (23 comparisons for seven models; appendix pp 8-9).

*Answer-extraction analysis.* Before a free-text reply can be scored, the chosen option letter must be extracted from it, and evaluation scripts do this in different ways. To measure the effect of this step alone, we re-scored all 20 300 stored multiple-choice replies of experiments 1–4, without new inference, with the platform’s final extraction rules, which produced every other score in this paper, and with four alternatives: an earlier version of the platform’s own rules; the regular expression of OpenAI simple-evals, which accepts only an explicit "Answer: X" line; and the first or the last option letter appearing in the reply, two simple rules used in evaluation scripts. Each alternative was compared with the final rules on the same replies (appendix pp 9–11).

*Experiment 2: protocol sensitivity.* Four models (Llama 3.1 8B, Qwen2.5 7B, Gemma 3 27B and Llama 3.3 70B) answered the same 150 MedQA questions under four protocols: the default prompt, an explicit instruction to answer with the option letter only, a chain-of-thought cue and three worked examples.

*Experiment 3: run-to-run stability.* Two models (Llama 3.1 8B and Gemma 3 27B) answered the same 150 MedQA questions three times at temperature 0 and three times at temperature 0·7.

*Experiment 4: image reliance.* Five vision-language models (Gemma 3 12B and 27B, Gemma 4 31B, Qwen3.8 27B and Qwen2.5-VL 7B) answered 150 questions from the five medical subjects of MMMU (basic medical science, clinical medicine, diagnostics and laboratory medicine, pharmacy and public health) and 150 from MedXpertQA MM (five options), each once with and once without the images.^24,26^

*Experiment 5: agentic computational biology.* Six models (Llama 3.1 8B, Qwen2.5 7B, Gemma 3 27B, Llama 3.3 70B, Gemma 4 31B and Qwen3.8 27B) were evaluated on two computational biology benchmarks. In BioCompute-Bench, which assesses single-step computational tasks, each model answered the same 75 questions under two conditions: without access to tools (closed-book) and with access to the sandboxed Python interpreter and bioinformatics sequence utilities. In BioAgent-Bench, which assesses multi-step analyses involving biological files, each model completed 40 tasks with access to the same tools.

### Statistical analysis

Accuracy is reported with Wilson 95% CIs. Pooled medical accuracy is question-weighted across the biomedical benchmarks. Because conditions were compared on identical items, paired differences between models, extraction rules, protocols, image conditions and tool conditions were tested with exact McNemar tests and reported with Newcombe hybrid score 95% CIs, with Holm correction within each experiment. In experiment 1, a published score was considered compatible with our measurement if it lay within the measured 95% CI; because the CIs of these comparisons had half-widths of 5·7–13·4 points (150 questions per benchmark, 50 per MMLU subset), compatibility does not establish equivalence, and results for individual MMLU subsets are descriptive. The number of questions needed to detect a paired protocol effect was estimated with the sample-size formula for McNemar’s test, using the proportion of discordant questions observed in experiment 2 (appendix p 15). In a sensitivity analysis, answers recovered from reasoning traces or obtained after truncation retries were excluded or counted as incorrect. Analyses used Python (pandas, SciPy) in notebooks that read the platform’s exports. Reporting follows the TRIPOD-LLM guideline where applicable;^27^ CONSORT-AI, SPIRIT-AI and DECIDE-AI do not apply because no trial or clinical deployment was involved. The study used only public benchmarks and involved no patients or personal data, so ethical approval was not required.

## Results

The five experiments comprised 205 runs (143 in experiment 1, 12 additional runs in experiment 2, 12 in experiment 3, 20 in experiment 4, and 18 in experiment 5) and 21 490 graded answers. All runs, including their prompts, model outputs, extracted answers and evaluation records, are accessible through the BenchmarkHub interface (Figure 1).

Across the 950 medical questions included in the pooled analysis, model accuracy ranged from 46·1% (95% CI 43·0–49·3) for Mistral 7B to 80·9% (78·3–83·3) for Gemma 4 31B (Figure 2). Qwen3.8 27B and Llama 3.3 70B achieved the next highest accuracies, at 75·8% (73·0–78·4) and 70·6% (67·7–73·4), respectively. Three of the four highest-ranked models were reasoning models, although performance varied substantially within this group: DeepSeek-R1 7B ranked tenth, with an accuracy of 47·5%. Reasoning models generated 4–17 times more tokens per question than instruction models (appendix p 13). Performance was particularly variable across individual benchmarks; accuracy on MedXpertQA Text ranged from 9% to 49% across models. After Holm correction for multiple comparisons, 47 of the 55 pairwise model comparisons showed statistically significant differences, including the comparison between the two highest-ranked models (appendix p 12). A sensitivity analysis excluding the 233 answers that were recovered from reasoning traces or obtained after truncation retries increased GLM-4.7-Flash’s pooled accuracy by 7·4 percentage points, moving it to second place. The remaining models changed by no more than 1·2 percentage points. Counting these answers as incorrect instead changed only the relative ranking of the two lowest-performing models (appendix p 14).

**Figure 2:**
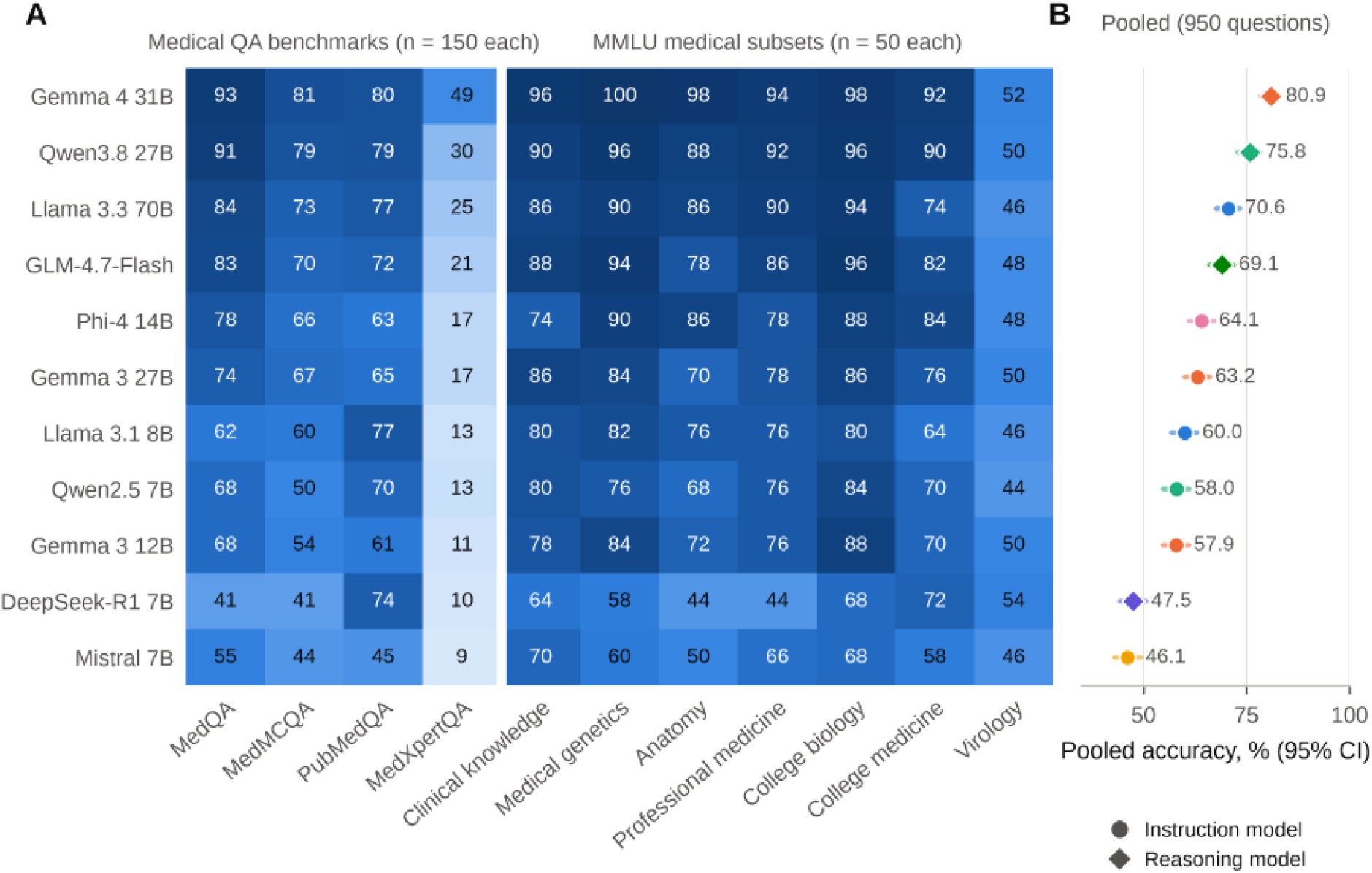
Accuracy of 11 locally served open-weight models on medical benchmarks. (A) Accuracy (%) per benchmark on the same seeded questions (150 per medical question-answering benchmark, 50 per MMLU subset). (B) Pooled, question-weighted accuracy over all 950 questions with Wilson 95% CIs; diamonds denote models that generate a reasoning trace. Rows are ordered by pooled accuracy.

Published scores were available for 23 model–benchmark pairs from seven models (appendix pp 8–9). Accuracy measured with the final rules was lower than the published value in 19 of them figure 3). Seven published values lay above the upper limit of the measured 95% CI, four of them on MMLU-Pro, for which developers reported five-shot chain-of-thought prompting, internal frameworks or incompletely specified settings. The other 16 lay within the measured CI and were therefore compatible with our measurements, although this does not establish equivalence.

**Figure 3:**
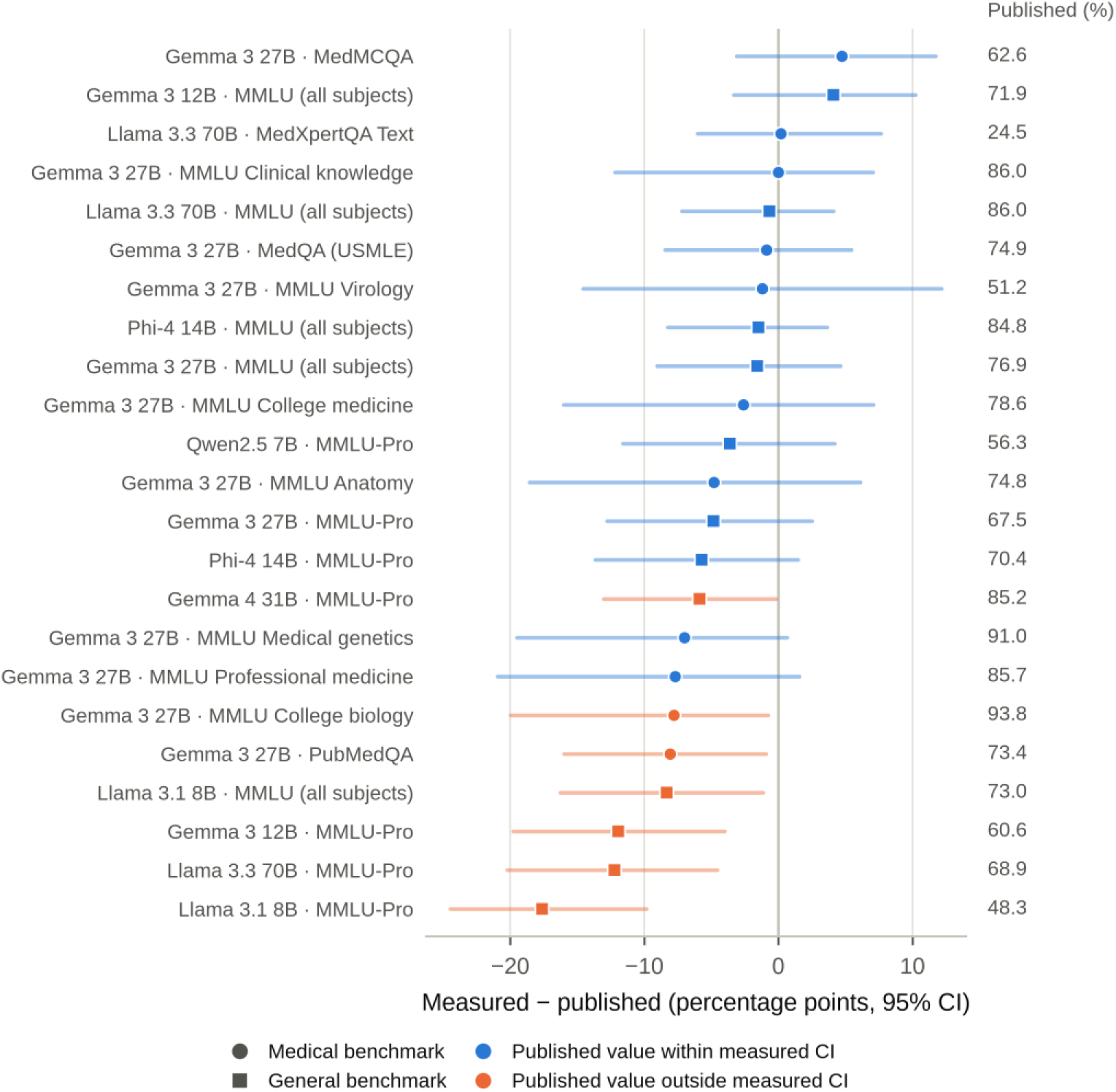
Agreement between published and measured scores. Difference between the accuracy measured in BenchmarkHub and the score published by the model developer or benchmark authors for 23 model–benchmark pairs; the published value is shown on the right. Bars are the 95% CI of the measured accuracy, expressed relative to the published value. Orange marks published values outside the measured CI; circles are medical and squares general benchmarks. A published value inside the CI is compatible with, but not shown to equal, the measured accuracy.

Evaluation choices changed measured accuracy by amounts comparable to the differences between models. The answer-extraction method had a substantial effect on measured accuracy, even though all methods were applied to exactly the same stored model replies (Figure 4A). Compared with the final BenchmarkHub extraction rules, selecting the first standalone option letter reduced pooled medical accuracy by up to 14·0 percentage points, for Gemma 4 31B. Selecting the last standalone option letter produced even larger accuracy reductions, reaching 33·1 percentage points for Gemma 3 27B. Compared with the final rules on the same replies, these reductions were statistically significant after Holm correction for five and nine of the 11 models, respectively. Both extraction methods also changed the relative ranking of models: with the first-letter rule, Qwen3.8 27B ranked first, whereas with the last-letter rule, Llama 3.3 70B ranked first and Gemma 3 27B fell to tenth place (appendix p 10). The OpenAI simple-evals pattern, which searches for an explicit “Answer: X” statement, identified only 1–48% of replies as answerable, depending on the model. Even the earlier and final versions of BenchmarkHub’s own rules graded 997 of the 20 300 stored replies (4·9%) differently, changing pooled accuracy by up to 4·3 percentage points (Gemma 4 31B) and the rank of six models (appendix p0 10-11).

**Figure 4:**
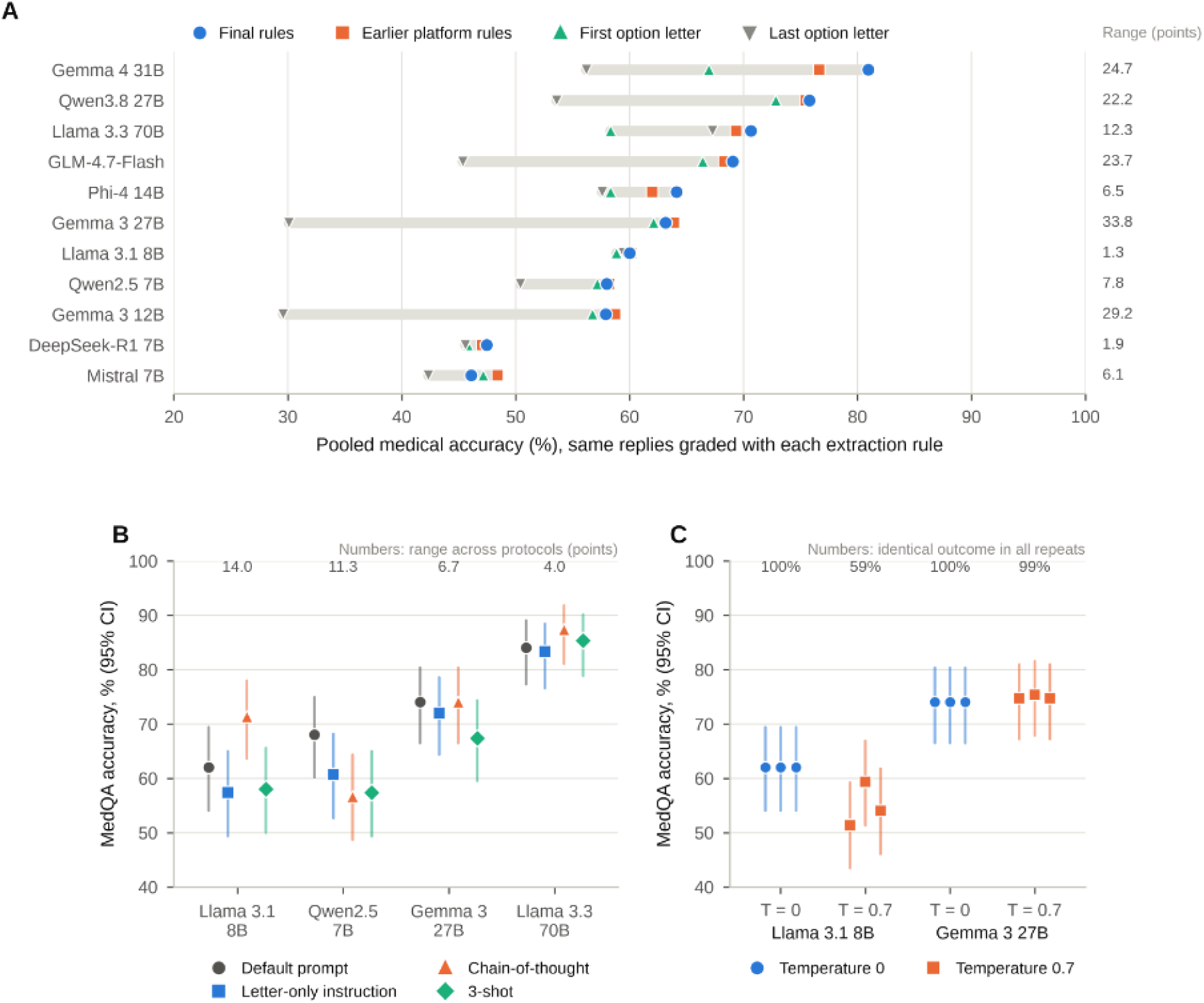
Effect of evaluation choices on measured accuracy. (A) Pooled medical accuracy (950 questions per model) of the same stored replies graded with the platform’s final extraction rules, an earlier version of its rules, and the first or last standalone option letter; numbers give the range across rules in percentage points. (B) MedQA accuracy (Wilson 95% CI) of four models on the same 150 questions under four prompting protocols; numbers give the range across protocols. (C) MedQA accuracy (Wilson 95% CI) in three repeated runs at temperature 0 and 0·7; numbers give the percentage of questions with the same outcome in all three repeats. At temperature 0 the three repeats coincide.

In experiment 2, changing the prompting protocol altered MedQA accuracy by −11·3 to +9·3 percentage points relative to the default prompt (figure 4B; appendix p 14-15). The direction and magnitude of these changes depended on the model. A chain-of-thought cue increased accuracy for Llama 3.1 8B by 9·3 percentage points (95% CI 1·0–17·5), but decreased accuracy for Qwen2.5 7B by 11·3 percentage points (4·0–18·4). However, none of the 12 protocol comparisons remained statistically significant after Holm correction; the smallest adjusted p value was 0·055. Approximately 19% of questions changed from correct to incorrect, or vice versa, between protocols. Based on this observed discordance, approximately 600 questions would be required to detect a 5-percentage-point protocol effect with 80% power. In experiment 3, repeated runs at temperature 0 produced identical results for both models. At temperature 0·7, however, run-to-run variability differed between models. Llama 3.1 8B achieved accuracies ranging from 51·3% to 59·3% across the three runs (SD 4·1 percentage points), with identical outcomes across all three runs for only 59·3% of questions. Gemma 3 27B was substantially more stable, with accuracy ranging from 74·7% to 75·3% and identical outcomes across all three runs for 98·7% of questions (figure 4C; appendix p 15).

In the image-ablation experiment on multimodal benchmarks (experiment 4), removing images from the input lowered accuracy on the medical subjects of MMMU for all five vision-language models, although the magnitude of the reduction varied (figure 5; appendix p 15). The largest reductions were observed for Qwen3.8 27B, whose accuracy decreased by 19·3 percentage points (11·7–26·7), and Gemma 4 31B, whose accuracy decreased by 18·0 percentage points (95% CI 10·0–25·6). Both differences remained statistically significant after Holm correction (adjusted p<0·001). For the other three models, removing images reduced accuracy by 6·7–9·3 percentage points, but these differences were not statistically significant after correction. On MedXpertQA MM, adding the image did not detectably improve accuracy for any model (differences −4·0 to +6·0 percentage points, all non-significant); for the two weakest models accuracy was even nominally lower with the image. However, the corresponding CIs did not exclude potential accuracy gains of up to 15 percentage points from image availability (appendix p 15). Notably, the two highest-performing models answered 49·3% and 50·0% of MedXpertQA MM questions correctly even without access to the images, compared with 55·3% and 56·0%, respectively, when images were provided. Both text-only accuracies were above the 20% chance level for this five-option benchmark. Gemma 4 31B’s developer-reported accuracy on MedXpertQA MM was 61·3%, compared with 55·3% measured here with images.

**Figure 5:**
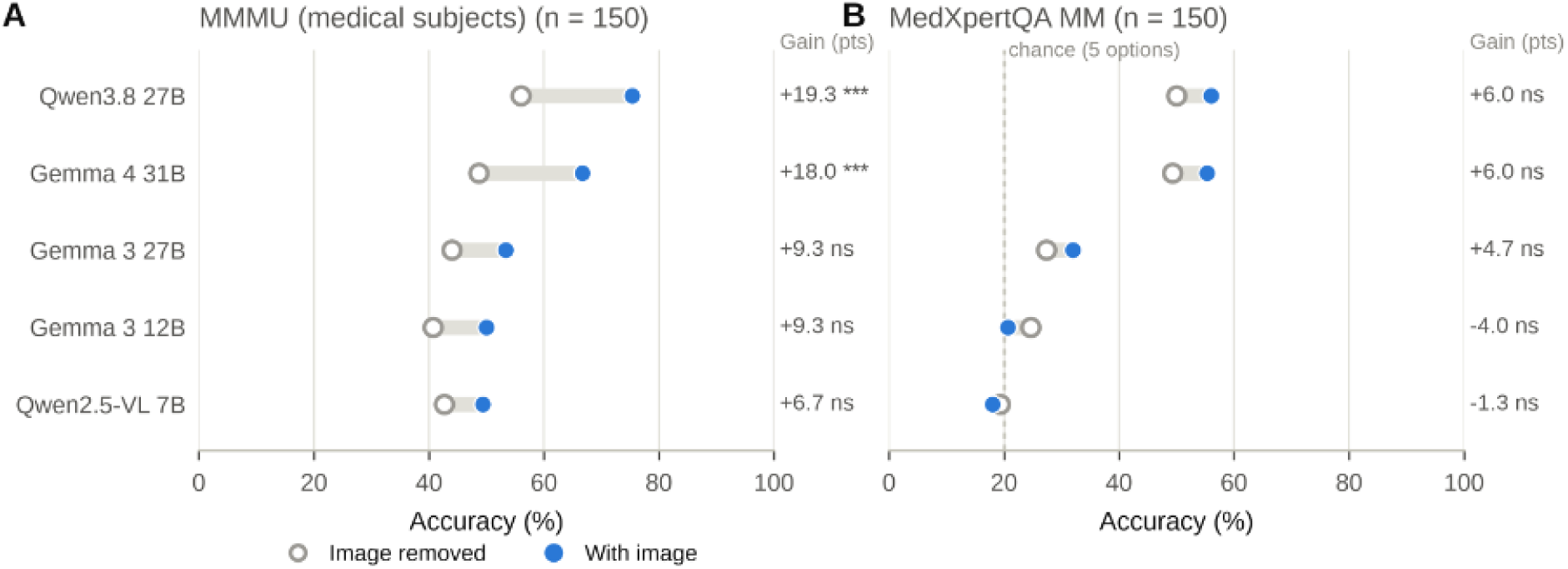
Image reliance on medical multimodal benchmarks. Accuracy of five vision-language models with the image and with the image removed, on identical questions from (A) the five medical subjects of MMMU and (B) MedXpertQA MM. Numbers give the gain from the image (accuracy with minus without image, percentage points) and the Holm-adjusted significance of exact McNemar tests (\*\**\** p<0·001; ns, not significant). The dashed line marks chance accuracy on MedXpertQA MM.

In the tool-ablation experiment on computational biology benchmarks (experiment 5), access to the sandbox and sequence-analysis tools substantially improved performance on BioCompute-Bench (figure 6A). Without tools, accuracy across the six models ranged from 4·0% to 56·0%, with Qwen3.8 27B achieving the highest score. When the same 75 items were evaluated with access to the sandbox and sequence tools, accuracy increased to between 41·3% and 98·7%. The paired improvements ranged from 36·0 percentage points (95% CI 20·5–49·2) to 62·7 percentage points (48·6–72·9), and all comparisons were statistically significant after Holm correction (p<0·001). The largest improvements were observed in task families requiring character-level sequence manipulation, such as reverse-complement generation and translation, which models rarely solved without tools but usually solved when the relevant tools were available (figure 6B).

**Figure 6:**
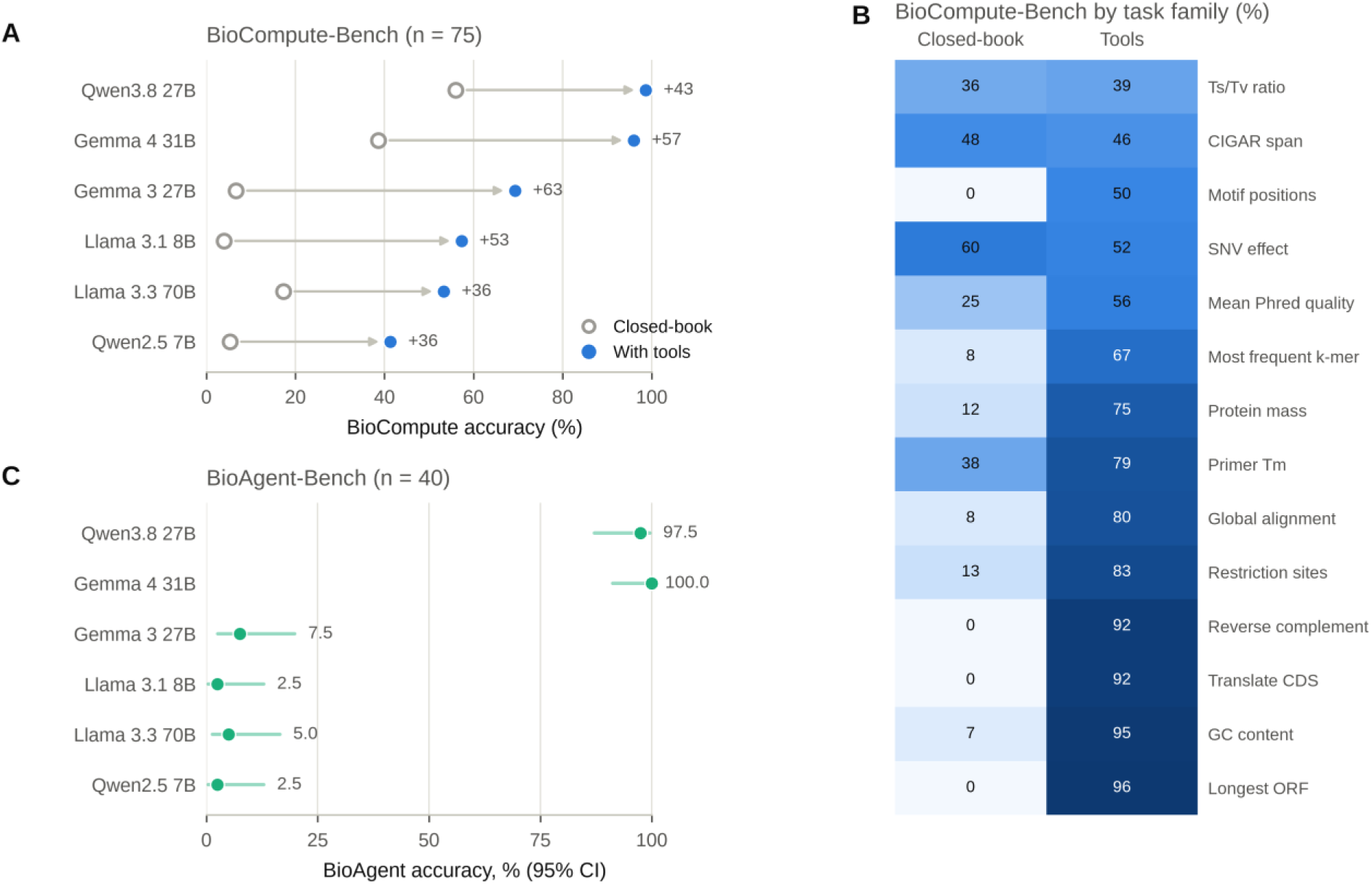
Agentic computational biology. (A) BioCompute-Bench accuracy on the same 75 items closed-book and with a sandboxed Python interpreter and sequence tools; numbers give the gain in percentage points. (B) BioCompute-Bench accuracy per task family under each condition, pooled over the six models; the 75-item sample contained 14 of the 15 families (no Hamming distance items). (C) BioAgent-Bench accuracy (95% CI) on 40 multi-step analyses of generated FASTA, VCF, GFF3, FASTQ and expression files, with tools.

Performance on the multi-step BioAgent-Bench tasks varied markedly between models (figure 6C). Gemma 4 31B solved all 40 tasks (100·0%), and Qwen3.8 27B solved 39 (97·5%). In contrast, Llama 3.1 8B and Qwen2.5 7B each solved one task (2·5%), while Llama 3.3 70B solved two (5·0%). Tool-trajectory analysis showed that successful multi-step analyses generally required the Python interpreter: 83 of the 86 correctly solved model–task instances involved Python. By comparison, only one of the 25 tasks attempted using sequence tools alone was solved. For single-step BioCompute-Bench tasks, sequence tools alone were used successfully in a similar proportion of the tasks in which they were invoked as Python (83% versus 80%, respectively; appendix pp 16-17). The recorded trajectories also revealed different failure patterns. Llama 3.3 70B typically stopped after a single tool call, whereas Qwen2.5 7B made a mean of 48 tool calls per task, most of them to sequence utilities, with frequent errors. Finally, independent recomputation reproduced the stored ground-truth answers for all 380 procedurally generated benchmark items (appendix p 8).

## Discussion

BenchmarkHub makes model evaluation a recorded, repeatable and locally executable procedure. Using it on 12 open-weight models, we found that the answer-extraction rule alone changed medical accuracy by more than the differences between many models and reordered the leaderboard, that published scores were usually higher than those measured under a documented protocol, and that image availability and tool access changed results substantially, whereas prompting-protocol effects of up to 11 points could not be estimated precisely with 150 questions. A single number on a model card therefore says little unless the procedure that produced it is reported and can be rerun.

Answer extraction is usually treated as an implementation detail, but it was the largest source of variation among the evaluation settings we examined. Sensitivity to prompt format and answer extraction has been documented on general tasks,^14–16^ and harness developers name extraction and normalisation as recurrent sources of irreproducibility;^18^ we show that on medical benchmarks simple rules found in evaluation scripts change accuracy by up to 33 points for the same replies, penalising models for their reply style rather than their knowledge, and that even careful revisions of a rule move scores and ranks. Because the platform keeps every raw reply, extraction rules can be compared and audited after the fact, and all runs re-scored without new inference. Medical evaluations should report their extraction rules and, ideally, release raw replies.

The gap with published scores, largest on MMLU-Pro, is consistent with those scores having been obtained with different prompting protocols and internal frameworks; quantisation is an unlikely sole explanation, because the largest gap was for the only model served with 16-bit weights. It illustrates why published and locally measured numbers should not be compared without their protocols. Typical medical evaluation sizes are also too small to estimate protocol effects: with 150 questions, differences of about 10 points between prompts were compatible with chance, and several hundred questions per condition are needed. Temperature 0 gave identical repeats in our setting, in which one local server processed requests without batching, but it does not guarantee reproducibility in general: when models are run through APIs or with batched inference, outputs at temperature 0 still vary between runs,^17,28^ so serving conditions belong in the methods of any evaluation and repeated runs are advisable.

The image ablation shows that multimodal accuracy can overstate visual understanding. Text-only shortcuts have been documented for general multimodal benchmarks^19^ and flawed visual reasoning behind correct answers for medical image challenges.^20^ MedXpertQA MM was filtered for difficulty rather than for image necessity,^24^ and its original report did not include a text-only baseline; the two strongest models answered half of its items correctly without the image, and no model gained significantly from it. On MMMU^26^ medical subjects, by contrast, the strongest models clearly used the image. Running every vision benchmark with and without images is a cheap control that the platform performs on identical items.

The agentic experiment adds a dimension that static question answering misses. Biology benchmarks without tools show that LLMs handle sequence manipulation poorly,^29^ and agentic benchmarks over real analyses remain far from solved.^30^ On our procedurally generated tasks, a sandboxed interpreter converted near-zero closed-book accuracy on character-level computations into near-perfect accuracy for the larger models, and multi-step analyses depended on writing code rather than on the predefined sequence tools, whereas small models stopped early or looped through failing tool calls. Procedural generation with computed, independently verified ground truth makes these benchmarks exactly gradable and resistant to memorisation, and the stored trajectories make agent failures diagnosable.

For health systems, running open-weight models on premises is attractive for data protection but shifts the burden of evaluation to the institution. BenchmarkHub lets a hospital or research group evaluate models on public benchmarks and its own questions without data leaving its infrastructure, compare them under identical documented conditions with paired statistics, and export complete records for audit, as recommended by reporting guidance for LLM studies.^27^ Such evaluations are a screening step: benchmark accuracy is not clinical utility,^7–9^ and our results add that even benchmark accuracy is conditional on the evaluation procedure. The platform can also host clinician-defined task suites^11^ and new clinical benchmarks.^12,13^

This study has limitations. Benchmarks were sampled (150 questions per benchmark, 50 per MMLU subset), so individual accuracies carry CIs of about ±6–8 points (±10–14 for MMLU subsets) and protocol effects were underpowered; the platform can run complete benchmarks. The final extraction rules were refined on replies from the same models and may be less accurate for other models. All but one model were 4-bit builds, which may explain part of the differences from published scores, and the published scores available were few and heterogeneous in protocol. The generated benchmarks cover well defined computational tasks rather than open research questions. The sandbox limits resources and blocks network access but does not isolate the filesystem or filter system calls, so untrusted models require a container or virtual machine (appendix p 6). Finally, all benchmarks were in English and none used real patient records.

In conclusion, medical benchmark results depend on how they are produced. Open, self-hosted evaluation infrastructure that records each step allows institutions to reproduce, audit and extend claims about LLMs before considering them for clinical use.

### Contributors

JIA-A conceived and developed the software, designed and ran the experiments, analysed the data and wrote the first draft. DJ-C and FS-C supervised the work, contributed to study design and interpretation and revised the manuscript. All authors had full access to all the data and had final responsibility for the decision to submit for publication.

### Data sharing

The BenchmarkHub source code is available under the MIT licence at https://github.com/juanjuanignacio/benchmarkhub. The experiment plan, client scripts, exported run records (every question, raw reply, extracted answer, correctness and agent trajectory), analysis notebooks and figure sources are available in the paper_experiments directory of the same repository from the date of publication, without restriction. All benchmarks used are publicly available from their original sources; the procedurally generated benchmarks are regenerated identically by the software.

### Declaration of generative AI use

During the development of the BenchmarkHub software, the authors used Claude Code (Anthropic; Opus 5.5) to assist with writing and revising code. During the preparation of this manuscript, the authors used the same tool to develop a draft written by the authors into the submitted text. The study design, the choice of models and benchmarks, the execution of the experiments, the analysis and interpretation of the results, and all decisions about the content of the article were made by the authors. The authors reviewed, edited and verified all AI-assisted code and text, and take full responsibility for the content of the publication.

## Data Availability

All data produced are available online at https://github.com/juanjuanignacio/benchmarkhub

## Acknowledgments

This work was supported by the Joint Action on Cardiovascular Diseases and Diabetes (JACARDI), which has received funding from the EU4Health Programme 2021–2027 under grant agreement 101126953; by grant PID2022-141527OB-I00 funded by MCIN/AEI/10.13039/501100011033 and by FEDER, UE; by the European Union Recovery and Resilience Facility–NextGenerationEU, within the call for R&D Projects of Excellence in Artificial Intelligence of the Secretary of State for Digitalisation and Artificial Intelligence; and by NextGenerationEU funds under the Recovery, Transformation and Resilience Plan through the talent attraction and retention programmes (Cardiotraining). The CNIC is supported by the Instituto de Salud Carlos III (ISCIII), the Ministerio de Ciencia, Innovación y Universidades (MICIU) and the Pro CNIC Foundation, and is a Severo Ochoa Center of Excellence (grant CEX2020-001041-S funded by MICIU/AEI/10.13039/501100011033). The views and opinions expressed are those of the authors only and do not necessarily reflect those of the European Union or the European Health and Digital Executive Agency (HaDEA). Neither the European Union nor the granting authority can be held responsible for them.

---

**Table 1:** Experiments run through BenchmarkHub.

| Experiment | Question | Models | Benchmarks and items | Conditions | Analysis |
|---|---|---|---|---|---|
| 1 Medical leaderboard | How accurate are locally served open models, and are published scores reproduced? | 11 text models | MedQA, MedMCQA, PubMedQA, MedXpertQA Text (150 each); seven MMLU medical subsets (50 each); MMLU, MMLU-Pro (150 each) | Zero-shot, temperature 0 | Wilson CI; McNemar (Holm); 23 published scores; sensitivity analysis |
| Answer extraction (re-analysis) | How much does the extraction rule change a score? | 12 models (11 text and 4 of them with image input, plus Qwen2.5-VL 7B) | All 20 300 stored multiple-choice replies of experiments 1–4 | Final rules; earlier platform rules; simple-evals pattern; first and last option letter | Paired differences (Newcombe CI), McNemar (Holm) |
| 2 Protocol sensitivity | How much does the prompting protocol change a medical score? | 4 | MedQA (150) | Default; letter-only instruction; chain-of-thought; three-shot | Paired differences vs default (Newcombe CI), McNemar (Holm) |
| 3 Stability | How variable are repeated runs? | 2 | MedQA (150) | Temperature 0 and 0·7, three repeats each | SD; identical outcomes |
| 4 Image reliance | Do vision-language models use the image? | 5 vision-language models | MMMU medical subjects (150); MedXpertQA MM (150) | With and without images | Paired differences (Newcombe CI), McNemar (Holm) |
| 5 Agentic biology | Do tools enable computational biology? | 6 | BioCompute-Bench (75); BioAgent-Bench (40) | Closed-book vs sandbox and sequence tools | Paired differences (Newcombe CI), McNemar (Holm); tool use from trajectories; independent ground truth |

All runs used seed 2026 for question sampling, so every model answered identical items. Full model, prompt and inference settings are in the appendix.

