## Supplemental Appendix for "BenchmarkHub: an open-source platform for reproducible evaluation of large language models in healthcare"

##### Contents

1. Models and inference settings
2. Benchmarks, sampling and prompts
3. Answer extraction and scoring rules
4. Agent loop and code sandbox
5. Procedurally generated benchmarks
6. Published scores used for comparison
7. Answer-extraction analysis
8. Supplementary results
9. Reproducing the study
Appendix references

#### 1. Models and inference settings

| Model | Ollama tag | Build digest (SHA-256, first 12) | Parameters (billions) | Weights | Reasoning trace | Image input | Reply limit (tokens) |
|---|---|---|---|---|---|---|---|
| Llama 3.1 8B | `llama3.1:8b-instruct-fp16` | `4aacac419454` | 8·0 | FP16 (16-bit) | No | No | 2048 |
| Mistral 7B | `mistral:latest` | `6577803aa9a0` | 7·2 | Q4_K_M (4-bit) | No | No | 2048 |
| Qwen2.5 7B | `qwen2.5:7b` | `845dbda0ea48` | 7·6 | Q4_K_M (4-bit) | No | No | 2048 |
| Phi-4 14B | `phi4:latest` | `ac896e5b8b34` | 14·7 | Q4_K_M (4-bit) | No | No | 2048 |
| Gemma 3 12B | `gemma3:12b` | `f4031aab637d` | 12·2 | Q4_K_M (4-bit) | No | Yes | 2048 |
| Gemma 3 27B | `gemma3:27b` | `a418f5838eaf` | 27·4 | Q4_K_M (4-bit) | No | Yes | 2048 |
| Llama 3.3 70B | `llama3.3:latest` | `a6eb4748fd29` | 70·6 | Q4_K_M (4-bit) | No | No | 2048 |
| DeepSeek-R1 7B | `deepseek-r1:7b` | `755ced02ce7b` | 7·6 | Q4_K_M (4-bit) | Yes | No | 8192 |
| GLM-4.7-Flash | `glm-4.7-flash:latest` | `4475827791a2` | 29·9 | Q4_K_M (4-bit) | Yes | No | 8192 |
| Gemma 4 31B | `gemma4:31b` | `6316f0629137` | 31·3 | Q4_K_M (4-bit) | Yes | Yes | 8192 |
| Qwen3.8 27B | `qwen3.8:27b` | `22130167c4c2` | 27·3 | Q4_K_M (4-bit) | Yes | Yes | 8192 |
| Qwen2.5-VL 7B | `qwen2.5vl:7b` | `5ced39dfa4ba` | 8·3 | Q4_K_M (4-bit) | No | Yes | 2048 |

**Model selection.** We chose open-weight models that could be served on a single institutional server through Ollama and that together cover the main open model families (Llama, Gemma, Qwen, Mistral, Phi, DeepSeek and GLM), sizes from 7 to 70 billion parameters, instruction and reasoning models, and, for the comparison with published scores, models whose developers or benchmark authors reported results on the benchmarks used. Vision-language models were those of this set that accept images, plus Qwen2.5-VL 7B.

Full build digests are in `paper_experiments/data/manifest/models.csv` (`bm_manifest`). All models were served by Ollama 0.32.14 on a shared multi-GPU server with a 32 768-token context window, temperature 0 (except experiment 3), at most three concurrent requests and a reply limit of 8192 tokens per turn in agentic runs.

#### 2. Benchmarks, sampling and prompts

| Benchmark (slug) | Source | Options | Items sampled | Prompt (platform default) |
|---|---|---|---|---|
| MedQA (`medqa_usmle`) | MedQA USMLE 4-option test set | 4 | 150 | "Question: {question}\nA) …\nB) …\nC) …\nD) …\nAnswer:" |
| MedMCQA (`medmcqa`) | MedMCQA validation set | 4 | 150 | As MedQA |
| PubMedQA (`pubmedqa`) | PubMedQA PQA-L (1000 expert-labelled items), full abstract | yes/no/maybe | 150 | "Abstract: {abstract}\n\nQuestion: {question}\nA) Yes\nB) No\nC) Maybe\nAnswer (A, B, or C):" |
| MedXpertQA Text (`medxpertqa_text`) | MedXpertQA Text test set (2450) | 10 | 150 | "{question with options}\n\nSelect the single best answer. End your reply with "Answer: <letter>" (A, …, J)." |
| MMLU medical subsets (7) | MMLU test set: clinical knowledge, medical genetics, anatomy, professional medicine, college biology, college medicine, virology | 4 | 50 each | As MedQA |
| MMLU (`mmlu`) | MMLU test set, all 57 subjects | 4 | 150 | As MedQA |
| MMLU-Pro (`mmlu_pro`) | MMLU-Pro test set | up to 10 | 150 | As MedQA with options A–J |
| MMMU (`mmmu`) | MMMU validation set; subjects Basic Medical Science, Clinical Medicine, Diagnostics and Laboratory Medicine, Pharmacy, Public Health | up to 4–9 | 150 | "{question, <image i> → [Image i]}\n\n(A) …\n(B) …\n\nAnswer with a single letter." |
| MedXpertQA MM (`medxpertqa_mm`) | MedXpertQA MM test set (2000 questions; images from the official archive) | 5 | 150 | As MedXpertQA Text |
| BioCompute-Bench (`biocompute`, `biocompute_agent`) | Procedurally generated (section 5) | typed answer | 75 | "{task}\n\nEnd your reply with a final line of the form:\nAnswer: <value>\nGive only the value on that line (no units, no explanation)." |
| BioAgent-Bench (`bioagent`) | Procedurally generated (section 5) | typed answer | 40 | As BioCompute-Bench, with task files in the sandbox working directory |

**Sampling.** Each run requested `num_questions` items with `sample_seed = 2026`; the platform draws `random.Random(2026).sample(sorted question ids, n)`, so every model receives the same items for a given benchmark. For MMMU the run additionally restricts the pool to the five medical subjects.

| Benchmark | Source (Hugging Face dataset) | Split | Revision | Items installed | Items evaluated | SHA-256 of evaluated items (first 16) |
|---|---|---|---|---|---|---|
| `bioagent` | generated, seed per item | – | platform code | 80 | 40 | `e00f8a9e9ae42338` |
| `biocompute` | generated, seed per item | – | platform code | 300 | 75 | `36c33110ae914848` |
| `biocompute_agent` | generated (same items as biocompute) | – | platform code | 300 | 75 | `df2f6fb049eadd77` |
| `medmcqa` | openlifescienceai/medmcqa | validation | `91c6572c4540` | 4 183 | 150 | `16cb66beccf9cfb9` |
| `medqa_usmle` | GBaker/MedQA-USMLE-4-options | test | `0fb93dd23a73` | 1 273 | 150 | `d1eb119a690dfc27` |
| `medxpertqa_mm` | TsinghuaC3I/MedXPertQA (MM) | test | `7e7c465a68eb` | 2 000 | 150 | `c987defe31198091` |
| `medxpertqa_text` | TsinghuaC3I/MedXPertQA (Text) | test | `7e7c465a68eb` | 2 450 | 150 | `129750c60b10317a` |
| `mmlu` | cais/mmlu (all) | test | `c30699e8356d` | 14 042 | 150 | `e89aec31047c4760` |
| `mmlu_anatomy` | cais/mmlu (anatomy) | test | `c30699e8356d` | 135 | 50 | `3af60efd95294572` |
| `mmlu_clinical_knowledge` | cais/mmlu (clinical_knowledge) | test | `c30699e8356d` | 265 | 50 | `64ff1bdff2b5bc45` |
| `mmlu_college_biology` | cais/mmlu (college_biology) | test | `c30699e8356d` | 144 | 50 | `009ddb3418b6327b` |
| `mmlu_college_medicine` | cais/mmlu (college_medicine) | test | `c30699e8356d` | 173 | 50 | `cdb8e400fb9ac1fc` |

| | | | | | | |
|---|---|---|---|---|---|---|
| mmlu_medical_genetics | cais/mmlu (medical_genetics) | test | c30699e8356d | 100 | 50 | a35100d0b4084bba |
| mmlu_pro | TIGER-Lab/MMLU-Pro | test | 527feea0afed | 12 032 | 150 | 9afbda12d469585c |
| mmlu_professional_medicine | cais/mmlu (professional_medicine) | test | c30699e8356d | 272 | 50 | c4af85f0d764ab2e |
| mmlu_virology | cais/mmlu (virology) | test | c30699e8356d | 166 | 50 | 84a65c89cc6de44b |
| mmmu | MMMU/MMMU (5 medical subjects) | validation | 4619a102cf5a* | 900 | 150 | 2ae92720f944a296 |
| pubmedqa | qiaojin/PubMedQA (pqa_labeled) | train | 9001f2853fb8 | 1 000 | 150 | 7b2b36eda885486d |

Revision: commit of the dataset repository in the Hugging Face cache used to load the benchmark (*datasets cache revision). The fingerprint is the SHA-256 over the sorted per-item digests of question, options, answer, context, metadata and image bytes (`apps/benchmarks/fingerprint.py`); full values for the installed and evaluated sets are in `paper_experiments/data/manifest/benchmarks.csv`. Another installation can confirm that it evaluates identical items with `python manage.py bm_manifest`.

**Protocols (experiment 2).** Default: the prompt above. Letter-only: the system prompt “Answer with only the letter of the correct option.” placed before the question. Chain-of-thought: the suffix “Let’s think step by step.” appended to the prompt. Three-shot: three examples drawn with the run seed from MedQA questions not in the evaluated set, each shown as its prompt followed by its correct letter, introduced by “Here are some example questions and answers:” and followed by “Now answer the following:”.

**Image ablation (experiment 4).** Runs with `drop_images = true` send the identical prompt without the image attachments; questions, order and grader are unchanged.

#### 3. Answer extraction and scoring rules

*The text of this section is identical to the “Answer Extraction and Scoring” section of the software documentation (README.md).*

All scores are computed by deterministic rules in code: there is no LLM judge and no partial credit. Every result stores the raw reply, the extracted answer (`parsed_answer`) and, when relevant, flags explaining how the answer was obtained, so any score can be audited from the run detail page, the CSV/Excel export or the API. When a grader is improved, `python manage.py bm_regrade` re-scores stored replies without new inference (`--tag`, `--benchmark` or run ids; `--dry-run` shows what would change).

##### 1. Reasoning traces are removed first

Before any extraction the reply is passed through `strip_reasoning` (`apps/benchmarks/loader_base.py`):

- `<think>…</think>`, `<thinking>…</thinking>` and `<reasoning>…</reasoning>` blocks are removed; if only a closing tag appears (the chat template opened it), everything before it is removed.
- A reply that opens a reasoning block and never closes it (truncated mid-thought) is treated as **empty**, never graded on the half-finished reasoning.
- Reasoning returned in a separate field (OpenAI/vLLM `reasoning_content`, Ollama `thinking`) is stored in the result metadata (`reasoning`) and is not graded.

##### 2. Truncated replies

Reasoning models can spend the whole `max_tokens` budget thinking. When a single-completion reply ends with `finish_reason = length` **and** has no visible answer, the runner retries once with the same prompt plus *"a previous attempt ran out of tokens while reasoning. Reason briefly and make sure to state your final answer."* Tokens and time of both attempts are summed and the result is flagged `truncation_retry`. If the answer is still empty but the backend returned a reasoning trace, the last 1,500 characters of that trace are graded and the result is flagged `answer_source: reasoning`. Both flags are counted per run in exports.

##### 3. Multiple-choice letters

Used by MMLU (all subsets), MedQA, MedMCQA, MedXpertQA, MMLU-Pro, AQuA-RAT, GPQA, MMMU and every loader that keeps the default `evaluate_answer`. `_parse_mc_answer(reply, letters)` only accepts the benchmark's valid letters (A–D, A–E, or A–J for 10-option sets) and works on the original casing, so the article "A" or the pronoun "I" is not read as an option. Rules are tried in order and the first that matches wins:

Before the rules are applied, parenthesised lists of options such as `(A, B, or C)` or `(A, B, C, D)` are removed: models often echo the prompt's own line (`Answer (A, B, or C): B`), and the list is never an answer.

| # | Rule | Example that matches |
|---|---|---|
| 1 | LaTeX box with a letter, or with the exact text of exactly one option | `\boxed{C}`, `\boxed{\text{C}}`; `\boxed{59}` when option A is `59` |
| 2 | Explicit answer statement — the **last** one | `The answer is (B)`, `Answer: d`, `final answer would be **C**` |
| 3 | Inverse statement — last one | `B is the correct answer`, `making D the best option` |
| 4 | Bold claim | `**B) Cutaneous xanthomas** are the most likely finding` |
| 5 | Closing line that states an option | last line `**D) Cefepime and levofloxacin**` or `…is **Methadone (C)**.` (exactly one `(X)` on the line) |
| 6 | First inline statement of an option | `The most likely diagnosis is **A) Sarcoidosis**`, `the best next step is (B)` — not `The options are: A) …` |
| 7 | First bold option anywhere | `**C) Hypertension**` |
| 8 | Leading lone letter | `B`, `B)`, `(B).` — not `A detailed analysis…` |
| 9 | First line formatted as an option | `…the answer is:\n\nA) Disclose the error` (bullets allowed) |
| 10 | Lone letter on the last line | `…\n\nC` |
| 11 | First standalone valid letter, skipping `A`/`I` used as words | `I think C fits best` → C |

If nothing matches, the extracted answer is empty and the question is scored wrong. The extracted letter is compared with the stored correct letter.

The order reflects how models actually answer: verbose models state their choice ("the diagnosis is **A) …**") and then discuss the other options, so a statement or a conclusion line outranks the first letter that happens to appear. These rules were validated on the stored replies from 11 open models (see `paper_experiments/README.md`) and each rule has regression tests built from real replies in `tests/test_agentic.py`.

#### 4. Label answers (yes/no/maybe, true/false, NLI)

PubMedQA (`yes`/`no`/`maybe` or A/B/C), BoolQ (`true`/`false`/`yes`/`no`) and MultiNLI (`entailment`/`neutral`/`contradiction` or A/B/C) use `_parse_label`, which matches **whole words only** ("no" is never found inside "not" or "known"). Echoed option lists (`Answer (A, B, or C): B`) are removed first, as for multiple choice. In order: the last explicit `answer: X`; the reply's first word (a single letter is only accepted there if it is a valid option letter, or upper-case); otherwise the first label word that appears anywhere.

#### 5. Other single-completion graders

| Benchmarks | Rule |
|---|---|
| GSM8K | Last number in the reply (thousands separators removed); correct if within 0.01 of the truth |
| MATH-500 | Final value from `\boxed{}`, then a trailing `= x`, then `answer is` / `Answer:`, then the last number; equal after LaTeX normalisation, numerically equal or present as a whole token |
| TriviaQA | Lower-cased, punctuation removed; correct if the answer or any official alias appears as a whole phrase |
| NQ-Open | Correct if any accepted answer appears in the lower-cased reply (substring) |
| SimpleQA | Correct if the lower-cased answer appears in the reply or the |

| | |
|---|---|
| | reply in the answer (substring) |
| ChartQA | Official *relaxed accuracy*: numbers within 5% of the truth, otherwise normalised exact match |
| MMMU open questions | Normalised exact match |
| LibriSpeech | Word error rate (`jiwer`) < 0.1 |

##### 6. Agentic and tool-using runs

In the tool loop (`apps/agents/loop.py`) the answer is the text inside `<final_answer>…</final_answer>`, or the model's last reply without a tool call (after removing reasoning). Then:

- **Typed answers** (BioCompute, BioAgent, HubAgent) are graded by `metadata.answer_type` — see How they are graded.
- **Multiple-choice** benchmarks run with tools (e.g. LAB-Bench agent variants) use the parser in §3 on the final answer.
- **BFCL** grades the emitted calls by AST, and **GAIA** uses a port of the official scorer.

The loop also guards against outputs that are not answers, and every intervention is written to the trajectory:

- A reply truncated mid-reasoning gets up to two nudges to act, then a final turn without tools.
- In the ReAct text protocol, a reply that has neither a tool call nor a final answer (for example a plan announced without acting) gets up to two protocol reminders.
- At the step limit, the model must answer without tools.
- An empty final answer falls back to the reasoning trace (`answer_source: reasoning`).

##### 7. Prompt protocols

These settings change the prompt, never the grader:

- **System prompt.** Always placed first.
- **Few-shot.** Few-shot examples are drawn from questions outside the evaluated set. When the run has a `sample_seed` they are drawn with that seed, so the run is reproducible. Each example is shown as `prompt + correct answer`, after *"Here are some example questions and answers"*.
- **Chain of thought.** Appends *"Let's think step by step."*
- **`drop_images`.** Sends a vision benchmark without its images (text-only ablation), with the same questions and grader.
- **`sample_seed`.** Picks a reproducible random subset of `num_questions` items. Every model given the same seed sees the same questions, so paired tests (McNemar) are valid.

#### 4. Agent loop and code sandbox

**Protocol selection.** With providers that support function calling (here: Ollama models that declare tool support) tools are passed as JSON schemas and the model's `tool_calls` are executed. Otherwise the loop uses a text protocol in which the model writes `<tool_call>{"name": …, "arguments": {…}}</tool_call>` blocks and gives its answer in `<final_answer>…</final_answer>`. In automatic mode the loop falls back to the text protocol when the server reports that a model does not support tools (Gemma 3 in this study).

**System prompt.** “You are an autonomous agent solving a task. Work step by step and use the available tools whenever they help you get an exact, verified result (computations, sequence manipulation, reading files). Do not guess values you can compute. When you are confident, give the final answer.” followed by the protocol-specific instructions for calling tools and formatting the final answer.

**Limits and interventions.** Maximum assistant turns: 8 for BioCompute-Bench with tools and 15 for BioAgent-Bench. A reply truncated by the token limit before acting receives up to two reminders to act briefly; in the text protocol, a reply that contains neither a tool call nor a final answer receives up to two protocol reminders; at the step limit the model is asked for its final answer without tools. Each intervention is recorded in the trajectory.

**Sandbox.** Python code runs in a separate interpreter started in isolated mode (`python -I`), with resource limits (CPU time 30 s, address space 2 GB, maximum file size 50 MB), inside a Linux user and network namespace without network access (`unshare -rn`), in a fresh temporary copy of the task's input directory that is deleted afterwards. Tool outputs longer than the observation limit are truncated before being returned to the model (6000 characters), and each call is stopped after 35 s.

**Limitations of the sandbox.** The sandbox limits resources and network access but is not a security boundary: code runs as the server user, so it can read files that user can read, and there is no filesystem isolation or system-call filtering. Untrusted models should be run inside a container or virtual machine, or with code execution disabled (`BENCHMARKHUB_SANDBOX=disabled`).

**Tools.** `python`, `calculator`, `list_files`, `read_file`, `seq_reverse_complement`, `seq_translate`, `seq_gc_content`, `seq_find_orfs`, `seq_restriction_sites`, `seq_align` (Needleman–Wunsch), `protein_mw`, `primer_tm`, plus `kb_search` and `kb_get` for the domain-neutral HubAgent-Bench (not used here). BioCompute-Bench with tools offers `python`, `calculator` and the sequence tools; BioAgent-Bench adds file listing and reading.

#### 5. Procedurally generated benchmarks

BioCompute-Bench, BioAgent-Bench and HubAgent-Bench were created for BenchmarkHub to fill two gaps: public bioinformatics test sets mostly measure *recall* (multiple-choice knowledge) rather than the ability to *compute* on sequences and files, and every public benchmark eventually leaks into training corpora. All three are **procedurally generated, exactly gradable and reproducible**.

##### How they are generated

1. **Families.** Each benchmark is a list of *task families* (15, 10 and 6 respectively). A family is one Python function, `generator(rng, qid, ctx)`, that writes a question of a given kind.
2. **Seeding.** For item *i* of family *f* the generator receives its own `random.Random(f"2026:{slug}:{f}:{i}")`. Every installation, on any machine and Python version, therefore produces **byte-identical questions and input files**, so scores are comparable across laboratories, while the items themselves never appear verbatim on the web.
3. **Random instances.** The generator draws a fresh instance: random DNA with a target GC content, coding sequences built from sense codons plus a stop codon, VCF records with realistic QUAL/FILTER/DP fields, multi-exon genes on both strands embedded in a random genome, expression matrices with replicate noise, fictional companies and people, web-server logs…
4. **Planted structure, unambiguous answers.** Where a question asks for "the" best item, the generator plants a clear winner and redraws until the answer is unique: a longest ORF that is strictly longest, a most frequent k-mer without ties, a differentially expressed gene 0.6 log2 units beyond the runner-up, a calendar that is guaranteed to contain a free slot. The test suite checks that every file-based task has a valid answer.
5. **Ground truth computed, never written by hand.** The answer is computed from the generated instance. For biology tasks this uses `apps/agents/bio.py`, **the same code that implements the agent's bioinformatics tools**, so the oracle and the tools can never disagree. The test suite regenerates BioCompute-Bench and checks that every stored answer passes its own grader and that generation is deterministic.
6. **Input files.** File-based tasks (BioAgent, HubAgent) write their inputs to `media/agent_tasks/<slug>/<question_id>/` when the benchmark is loaded. During a run, each question gets a fresh temporary copy of its folder as the sandbox working directory, so agents can't modify the originals or see other tasks.

##### BioCompute-Bench (`biocompute`, `biocompute_agent`) — 15 families × 20 = 300 items

Single-step computations that a bioinformatician would do with a script. Offered **closed-book** (`biocompute`) and **with tools** (`biocompute_agent`: Python sandbox + bio toolkit) on identical items: the difference measures how much a model gains from tools and whether it knows when to use them.

| Family | Example (abridged) | Answer |
|---|---|---|
| `reverse_complement` | Give the reverse complement (5'→3') of this DNA sequence: … | sequence |
| `translate_cds` | Translate this coding sequence into its protein using the standard genetic code… | sequence |
| `gc_content` | What is the GC content of this sequence, in percent, rounded to 2 decimals? | float |
| `longest_orf` | Considering both strands and all six reading frames, what is the length (aa) of the longest ATG-initiated ORF? | int |
| `hamming` | What is the Hamming distance between these two sequences? | int |
| `motif_positions` | List every 1-based start position (including overlaps) of the motif CCCTGC… | int list |
| `frequent_kmer` | Which 5-mer occurs most often (overlapping occurrences)? | sequence |
| `restriction_count` | How many XhoI recognition sites does this sequence contain? | int |
| `protein_mw` | Average molecular weight of this protein (Da), rounded to 1 decimal? | float |
| `primer_tm` | Melting temperature with $Tm = 64.9 + 41 \cdot (G+C-16.4)/N$, rounded to 1 decimal | float |
| `snv_effect` | A SNV changes position 24 from C to A. Synonymous, missense or nonsense? | label |
| `ts_tv_ratio` | Given these substitutions (A>C, G>A, …), what is the Ts/Tv ratio? | float |
| `cigar_span` | CIGAR 15M1I36M8D50M… — how many reference bases does the alignment span? | int |
| `mean_phred` | Mean Phred quality of this FASTQ quality string (Phred+33)? | float |
| `global_alignment` | Optimal Needleman–Wunsch score (match +1, mismatch −1, gap −2)? | int |

##### BioAgent-Bench (`bioagent`) — 10 families × 8 = 80 items

Multi-step analyses over **real file formats**. The agent has to list and read the files, write parsing and analysis code in the sandbox, and combine several steps. Tools: `python`, `calculator`, `list_files`, `read_file` and the sequence tools.

| Family | Input files | Task |
|---|---|---|
| `fasta_max_gc` | `contigs.fasta` (15–30 contigs) | Which contig has the highest GC content? |
| `fasta_longest_orf` | `transcripts.fasta` | Which record contains the longest ORF (both strands), and how long is it? |
| `vcf_titv` | `calls.vcf` (150–300 variants) | Ts/Tv ratio of PASS SNVs, indels excluded |
| `vcf_region_count` | `calls.vcf` | PASS indels with DP ≥ N inside a genomic interval |
| `expression_top_gene` | `counts.csv` (3 control + 3 treated) | Most up- or down-regulated gene by log2 fold change |
| `expression_deg_count` | `expression.tsv` | Number of genes passing fold-change and expression thresholds |
| `gff_cds_protein` | `genome.fasta` + `annotation.gff3` | Splice a multi-exon gene's CDS (respecting strand), translate it, report its length or last 10 residues |
| `fastq_quality` | `reads.fastq` | Reads with mean Phred ≥ threshold |
| `fastq_adapter` | `library.fastq` | Mean read length after adapter trimming |
| `plasmid_digest` | `plasmid.fasta` (circular) | Fragment sizes of a double digest, descending |

##### Independent validation of the ground truth

The answers of both benchmarks are computed by `apps/agents/bio.py`, the module that also implements the agent's sequence tools. `paper_experiments/validate_generated.py` recomputes every answer from the question text and input files with Biopython 1.88 and plain Python, without the platform code, and compares it with the stored answer using the item's tolerance.

All 380 of 380 items agreed.

| Benchmark | Task family | Items | Agreeing |
|---|---|---|---|
| biocompute | cigar span | 20 | 20 |
| biocompute | frequent kmer | 20 | 20 |
| biocompute | gc content | 20 | 20 |
| biocompute | global alignment | 20 | 20 |
| biocompute | hamming | 20 | 20 |
| biocompute | longest orf | 20 | 20 |
| biocompute | mean phred | 20 | 20 |
| biocompute | motif positions | 20 | 20 |
| biocompute | primer tm | 20 | 20 |
| biocompute | protein mw | 20 | 20 |
| biocompute | restriction count | 20 | 20 |
| biocompute | reverse complement | 20 | 20 |
| biocompute | snv effect | 20 | 20 |
| biocompute | translate cds | 20 | 20 |
| biocompute | ts tv ratio | 20 | 20 |
| bioagent | expression deg count | 8 | 8 |
| bioagent | expression top gene | 8 | 8 |
| bioagent | fasta longest orf | 8 | 8 |
| bioagent | fasta max gc | 8 | 8 |
| bioagent | fastq adapter | 8 | 8 |
| bioagent | fastq quality | 8 | 8 |
| bioagent | gff cds protein | 8 | 8 |
| bioagent | plasmid digest | 8 | 8 |
| bioagent | vcf region count | 8 | 8 |
| bioagent | vcf titv | 8 | 8 |

#### 6. Published scores used for comparison

| Model | Benchmark | Published (%) | Measured (%) (95% CI) | Source | Setting reported |
|---|---|---|---|---|---|
| Gemma 3 27B | MedQA | 74·9 | 74·0 (66·4–80·4) | MedGemma Technical Report (Table 3) [A1] | Computed internally by Google; MedQA 4-option test set; single inference, default temperature |
| Gemma 3 27B | MedMCQA | 62·6 | 67·3 (59·5–74·3) | MedGemma Technical Report (Table 3) [A1] | Computed internally by Google; single inference |
| Gemma 3 27B | PubMedQA | 73·4 | 65·3 (57·4–72·5) | MedGemma Technical Report (Table 3) [A1] | Computed internally by Google; single inference |
| Gemma 3 27B | MMLU anatomy | 74·8 | 70·0 (56·2–80·9) | MedGemma Technical Report (Table 3) [A1] | Computed internally by Google |
| Gemma 3 27B | MMLU clinical knowledge | 86·0 | 86·0 (73·8–93·0) | MedGemma Technical Report (Table 3) [A1] | Computed internally by Google |

| | | | | | |
|---|---|---|---|---|---|
| Gemma 3 27B | MMLU college biology | 93·8 | 86·0 (73·8–93·0) | MedGemma Technical Report (Table 3) [A1] | Computed internally by Google |
| Gemma 3 27B | MMLU college medicine | 78·6 | 76·0 (62·6–85·7) | MedGemma Technical Report (Table 3) [A1] | Computed internally by Google |
| Gemma 3 27B | MMLU medical genetics | 91·0 | 84·0 (71·5–91·7) | MedGemma Technical Report (Table 3) [A1] | Computed internally by Google |
| Gemma 3 27B | MMLU professional medicine | 85·7 | 78·0 (64·8–87·2) | MedGemma Technical Report (Table 3) [A1] | Computed internally by Google |
| Gemma 3 27B | MMLU virology | 51·2 | 50·0 (36·6–63·4) | MedGemma Technical Report (Table 3) [A1] | Computed internally by Google |
| Gemma 3 12B | MMLU | 71·9 | 76·0 (68·6–82·1) | Gemma 3 Technical Report (Table 18) [A2] | IT model, 0-shot |
| Gemma 3 12B | MMLU-Pro | 60·6 | 48·7 (40·8–56·6) | Gemma 3 Technical Report (Table 6) [A2] | IT model, 0-shot |
| Gemma 3 27B | MMLU | 76·9 | 75·3 (67·9–81·5) | Gemma 3 Technical Report (Table 18) [A2] | IT model, 0-shot |
| Gemma 3 27B | MMLU-Pro | 67·5 | 62·7 (54·7–70·0) | Gemma 3 Technical Report (Table 6) [A2] | IT model, 0-shot |
| Phi-4 14B | MMLU | 84·8 | 83·3 (76·5–88·5) | Phi-4 Technical Report (Table 1) [A3] | OpenAI simple-evals, temperature 0.5 |
| Phi-4 14B | MMLU-Pro | 70·4 | 64·7 (56·7–71·9) | Phi-4 Technical Report (Table 1) [A3] | Internal framework and answer extraction |
| Llama 3.1 8B | MMLU | 73·0 | 64·7 (56·7–71·9) | Llama 3.1 model card [A4] | 0-shot CoT, macro average |
| Llama 3.1 8B | MMLU-Pro | 48·3 | 30·7 (23·9–38·5) | Llama 3.1 model card [A4] | 5-shot CoT, macro average |
| Llama 3.3 70B | MMLU | 86·0 | 85·3 (78·8–90·1) | Llama 3.3 model card [A5] | 0-shot CoT, macro average |
| Llama 3.3 70B | MMLU-Pro | 68·9 | 56·7 (48·7–64·3) | Llama 3.3 model card [A5] | 5-shot CoT, macro average |
| Qwen2.5 7B | MMLU-Pro | 56·3 | 52·7 (44·7–60·5) | Qwen2.5 Technical Report (Table 8) [A6] | Instruct model |
| Gemma 4 31B | MMLU-Pro | 85·2 | 79·3 (72·2–85·0) | Gemma 4 31B model card [A7] | IT model; thinking setting not stated |
| Gemma 4 31B | MedXpertQA MM | 61·3 | not measured | Gemma 4 31B model card [A7] | IT model; thinking setting not stated |
| Llama 3.3 70B | MedXpertQA Text | 24·5 | 24·7 (18·5–32·1) | MedXpertQA paper (Table 5) [A8] | Zero-shot CoT, greedy decoding |

Only values verified in the primary source were used. The Gemma 4 MedXpertQA MM value is compared in experiment 4 (with-image condition). Model developers report few medical scores; most report only general benchmarks.

### 7. Answer-extraction analysis

#### Alternative extraction rules

Every stored multiple-choice reply of experiments 1–4 (20 300 replies; open-ended MMMU items excluded) was re-graded  without new inference  by `paper_experiments/extraction_audit.py` with five rules applied to the reply after removal of any reasoning trace:

- **Final rules**: the platform rules used for every score in the paper (section 3).
- **Earlier platform rules**: the rules of BenchmarkHub commit 9d03117, a previous version of those in section 3. They took the letter from a boxed answer, an explicit “answer is X” or “Answer: X”, a leading letter or a lone letter on the last line, and otherwise the first standalone capital letter (the only rule used for MMLU-Pro and MMMU); they did not recognise inline answer statements or conclusion lines, ignore option lists echoed from the prompt, or map boxed option values to letters, and PubMedQA labels were matched as substrings.
- **simple-evals regex**: the multiple-choice pattern of OpenAI simple-evals (“Answer: X”); it assumes a prompt that requests this format.

- **First option letter**: the first standalone letter among the valid options (`\b[A-J]\b`).
- **Last option letter**: the last standalone letter among the valid options.

| Model | Final rules | Earlier platform rules | First option letter | Last option letter | simple-evals regex | Range* (points) | Rank: final / earlier / first / last |
|---|---|---|---|---|---|---|---|
| Gemma 4 31B | 80·9 | 76·6 | 66·9 | 56·2 | 12·2 | 24·7 | 1 / 1 / 2 / 4 |
| Qwen3.8 27B | 75·8 | 75·5 | 72·8 | 53·6 | 6·1 | 22·2 | 2 / 2 / 1 / 5 |
| Llama 3.3 70B | 70·6 | 69·4 | 58·3 | 67·3 | 18·0 | 12·3 | 3 / 3 / 6 / 1 |
| GLM-4.7-Flash | 69·1 | 68·3 | 66·4 | 45·4 | 6·7 | 23·7 | 4 / 4 / 3 / 8 |
| Phi-4 14B | 64·1 | 62·0 | 58·3 | 57·6 | 3·5 | 6·5 | 5 / 6 / 6 / 3 |
| Gemma 3 27B | 63·2 | 63·9 | 62·1 | 30·1 | 2·6 | 33·8 | 6 / 5 / 4 / 10 |
| Llama 3.1 8B | 60·0 | 60·1 | 58·8 | 59·5 | 48·3 | 1·3 | 7 / 7 / 5 / 2 |
| Qwen2.5 7B | 58·0 | 58·2 | 57·2 | 50·4 | 2·6 | 7·8 | 8 / 9 / 8 / 6 |
| Gemma 3 12B | 57·9 | 58·7 | 56·7 | 29·6 | 1·8 | 29·2 | 9 / 8 / 9 / 11 |
| DeepSeek-R1 7B | 47·5 | 47·1 | 45·8 | 45·6 | 22·6 | 1·9 | 10 / 11 / 11 / 7 |
| Mistral 7B | 46·1 | 48·4 | 47·2 | 42·3 | 1·3 | 6·1 | 11 / 10 / 10 / 9 |

Pooled accuracy (%) on the 950 medical questions of experiment 1 for the same stored replies. *Range across the final, earlier, first-letter and last-letter rules.

| Rule compared with the final rules | Replies graded differently | Correct only under final rules | Correct only under the alternative |
|---|---|---|---|
| Earlier platform rules | 997 (4·9%) | 746 | 251 |
| First option letter | 1 961 (9·7%) | 1 580 | 381 |
| Last option letter | 3 464 (17·1%) | 3 016 | 448 |
| simple-evals regex | 10 188 (50·2%) | 10 177 | 11 |

All 20 300 stored multiple-choice replies of experiments 1–4.

| Model | Rule | Final minus alternative, points (95% CI) | Holm-adjusted p |
|---|---|---|---|
| DeepSeek-R1 7B | First option letter | 1·7 (0·5 to 2·8) | 0·11 |
| GLM-4.7-Flash | First option letter | 2·6 (1·1 to 4·1) | 0·02 |
| Gemma 3 12B | First option letter | 1·2 (0·2 to 2·1) | 0·45 |
| Gemma 3 27B | First option letter | 1·1 (0·0 to 2·1) | 0·78 |
| Gemma 4 31B | First option letter | 14·0 (11·5 to 16·5) | <0·0001 |
| Llama 3.1 8B | First option letter | 1·2 (0·0 to 2·3) | 0·78 |
| Llama 3.3 70B | First option letter | 12·3 (9·9 to 14·7) | <0·0001 |
| Mistral 7B | First option letter | -1·1 (-1·9 to -0·2) | 0·43 |
| Phi-4 14B | First option letter | 5·8 (3·6 to 8·0) | <0·0001 |
| Qwen2.5 7B | First option letter | 0·8 (-0·3 to 2·0) | 1·00 |
| Qwen3.8 27B | First option letter | 2·9 (1·7 to 4·3) | 0·0002 |
| DeepSeek-R1 7B | Last option letter | 1·9 (0·7 to 3·1) | 0·06 |
| GLM-4.7-Flash | Last option letter | 23·7 (20·5 to 26·7) | <0·0001 |
| Gemma 3 12B | Last option letter | 28·3 (24·4 to 32·1) | <0·0001 |
| Gemma 3 27B | Last option letter | 33·1 (29·3 to 36·6) | <0·0001 |
| Gemma 4 31B | Last option letter | 24·7 (21·7 to 27·7) | <0·0001 |
| Llama 3.1 8B | Last option letter | 0·5 (-0·1 to 1·1) | 1·00 |
| Llama 3.3 70B | Last option letter | 3·4 (1·9 to 4·8) | 0·0001 |
| Mistral 7B | Last option letter | 3·8 (2·1 to 5·5) | 0·0005 |

| | | | |
|---|---|---|---|
| Phi-4 14B | Last option letter | 6·5 (4·7 to 8·4) | <0·0001 |
| Qwen2.5 7B | Last option letter | 7·6 (5·1 to 10·0) | <0·0001 |
| Qwen3.8 27B | Last option letter | 22·2 (19·4 to 25·0) | <0·0001 |
| DeepSeek-R1 7B | Earlier platform rules | 0·4 (-0·2 to 1·1) | 1·00 |
| GLM-4.7-Flash | Earlier platform rules | 0·7 (-0·2 to 1·6) | 1·00 |
| Gemma 3 12B | Earlier platform rules | -0·8 (-1·9 to 0·2) | 1·00 |
| Gemma 3 27B | Earlier platform rules | -0·7 (-1·7 to 0·3) | 1·00 |
| Gemma 4 31B | Earlier platform rules | 4·3 (2·9 to 5·8) | <0·0001 |
| Llama 3.1 8B | Earlier platform rules | -0·1 (-0·6 to 0·4) | 1·00 |
| Llama 3.3 70B | Earlier platform rules | 1·3 (0·5 to 2·1) | 0·03 |
| Mistral 7B | Earlier platform rules | -2·3 (-3·7 to -1·0) | 0·02 |
| Phi-4 14B | Earlier platform rules | 2·1 (0·0 to 4·2) | 0·68 |
| Qwen2.5 7B | Earlier platform rules | -0·2 (-1·3 to 0·9) | 1·00 |
| Qwen3.8 27B | Earlier platform rules | 0·3 (-0·5 to 1·1) | 1·00 |

Paired differences on the 950 medical questions (Newcombe hybrid score CI; exact McNemar test, Holm correction across the 33 comparisons shown and the 11 simple-evals comparisons).

### 8. Supplementary results

#### Table S1: Accuracy (%) by model and benchmark (experiment 1)

| **Model** | **MMLU (all subjects)** | **Anatomy** | **Clinical knowledge** | **College biology** | **College medicine** | **Medical genetics** | **Professional medicine** | **Virology** | **MMLU-Pro** | **MedMCQA** | **MedQA (USMLE)** | **MedXpert QA Text** | **PubMed QA** |
|---|---|---|---|---|---|---|---|---|---|---|---|---|---|
| Gemma 4 31B | 96·0 | 98·0 | 96·0 | 98·0 | 92·0 | 100·0 | 94·0 | 52·0 | 79·3 | 80·7 | 93·3 | 48·7 | 80·0 |
| Qwen3.8 27B | 92·0 | 88·0 | 90·0 | 96·0 | 90·0 | 96·0 | 92·0 | 50·0 | 79·3 | 78·7 | 91·3 | 30·0 | 79·3 |
| Llama 3.3 70B | 85·3 | 86·0 | 86·0 | 94·0 | 74·0 | 90·0 | 90·0 | 46·0 | 56·7 | 73·3 | 84·0 | 24·7 | 76·7 |
| GLM-4.7-Flash | 84·7 | 78·0 | 88·0 | 96·0 | 82·0 | 94·0 | 86·0 | 48·0 | 65·3 | 70·0 | 83·3 | 21·3 | 72·0 |
| Phi-4 14B | 83·3 | 86·0 | 74·0 | 88·0 | 84·0 | 90·0 | 78·0 | 48·0 | 64·7 | 66·0 | 78·0 | 16·7 | 62·7 |
| Gemma 3 27B | 75·3 | 70·0 | 86·0 | 86·0 | 76·0 | 84·0 | 78·0 | 50·0 | 62·7 | 67·3 | 74·0 | 16·7 | 65·3 |
| Llama 3.1 8B | 64·7 | 76·0 | 80·0 | 80·0 | 64·0 | 82·0 | 76·0 | 46·0 | 30·7 | 60·0 | 62·0 | 12·7 | 77·3 |
| Qwen2.5 7B | 69·3 | 68·0 | 80·0 | 84·0 | 70·0 | 76·0 | 76·0 | 44·0 | 52·7 | 50·0 | 68·0 | 13·3 | 70·0 |
| Gemma 3 12B | 76·0 | 72·0 | 78·0 | 88·0 | 70·0 | 84·0 | 76·0 | 50·0 | 48·7 | 54·0 | 68·0 | 11·3 | 60·7 |
| DeepSeek-R1 7B | 59·3 | 44·0 | 64·0 | 68·0 | 72·0 | 58·0 | 44·0 | 54·0 | 48·0 | 40·7 | 41·3 | 10·0 | 74·0 |
| Mistral 7B | 58·7 | 50·0 | 70·0 | 68·0 | 58·0 | 60·0 | 66·0 | 46·0 | 23·3 | 44·0 | 54·7 | 8·7 | 45·3 |

MMLU subsets have 50 questions each (95% CI half-width about 10–14 percentage points) and are descriptive; the other benchmarks have 150 (about 6–8 points).

**Figure S1: Paired comparisons between models on the medical battery**

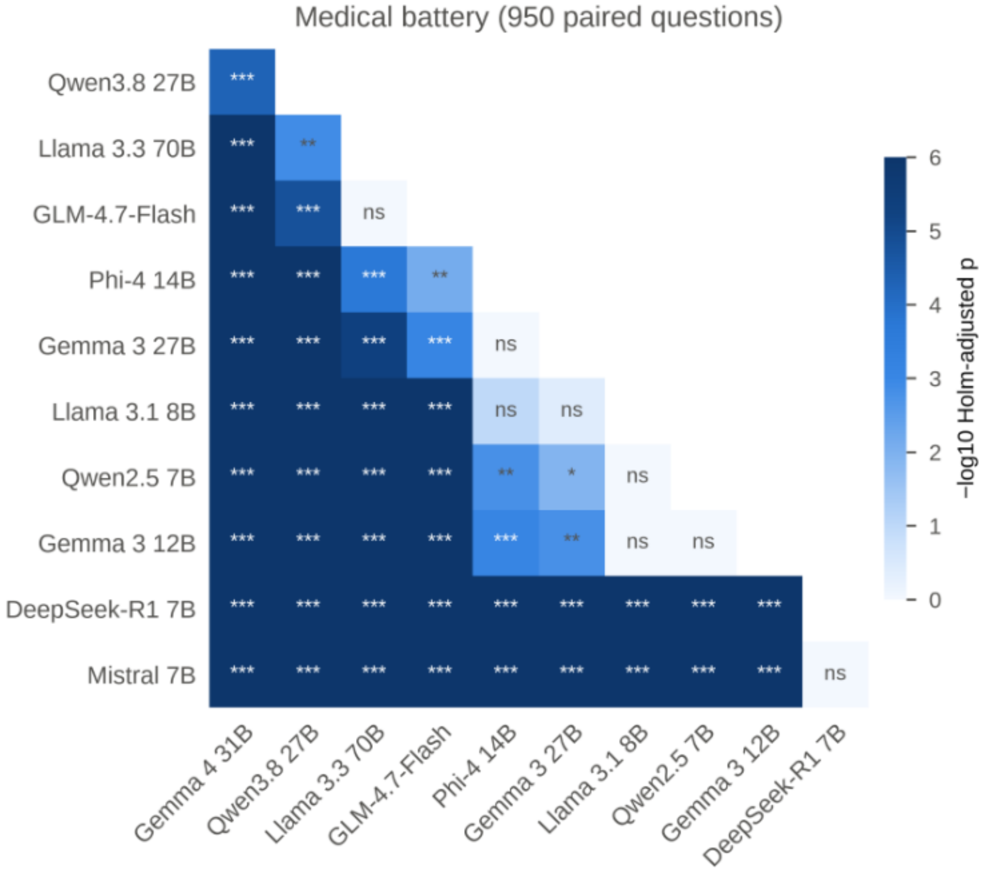


Exact McNemar tests on the 950 paired medical questions, Holm-adjusted across 55 model pairs (*** p<0·001, ** p<0·01, * p<0·05, ns not significant). Shade: −log10 adjusted p.

47 of 55 model pairs differed significantly after correction.

#### Figure S2: Paired comparisons on MedQA

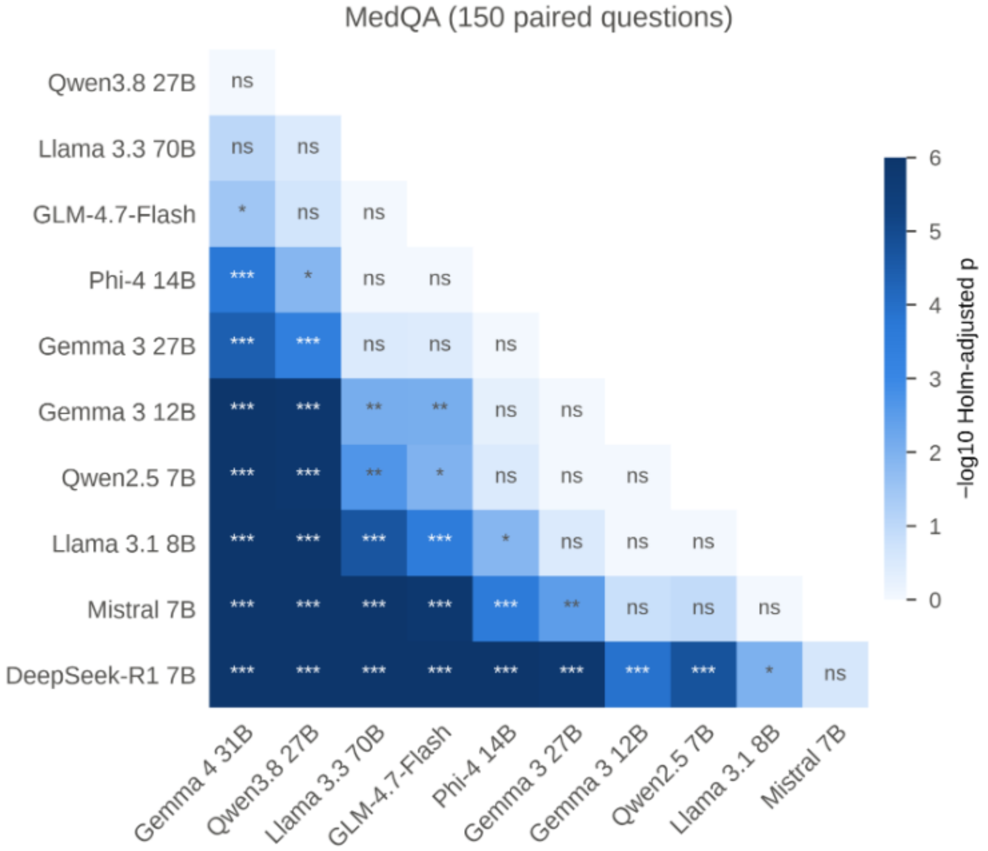


#### Figure S3: Accuracy versus inference cost

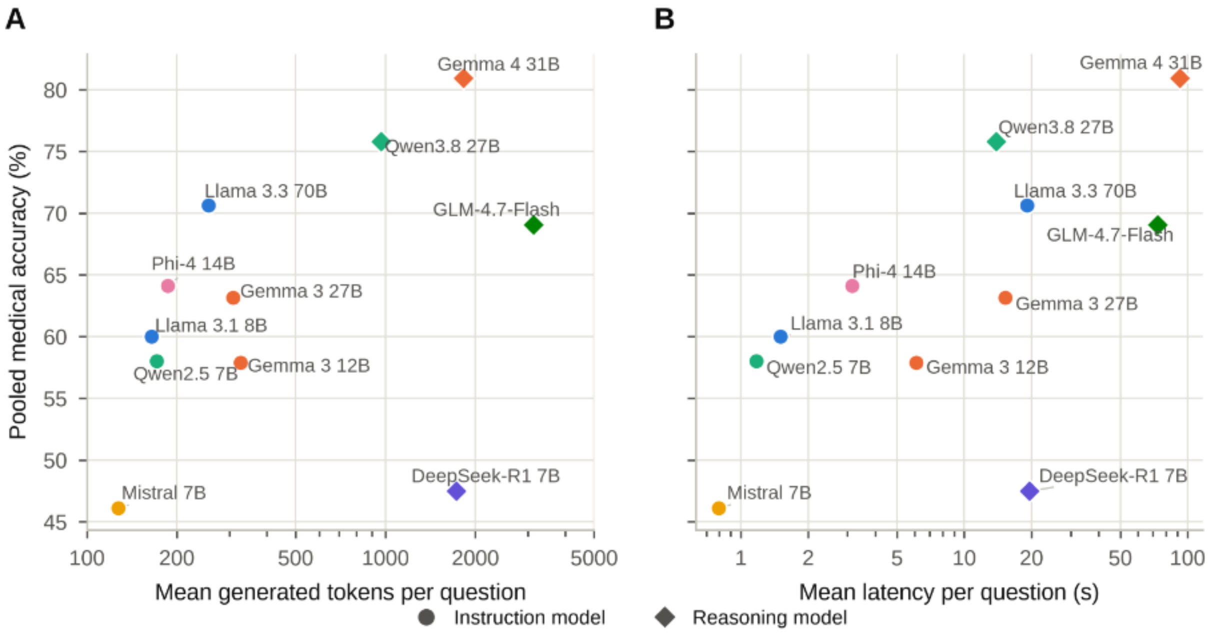


Pooled medical accuracy against (A) mean generated tokens per question and (B) mean wall-clock latency per question. Latency was measured on a shared server and is indicative only.

| Model | Pooled accuracy (%) | Mean latency (s) | Mean generated tokens |
|---|---|---|---|
| Gemma 4 31B | 80·9 | 92·2 | 1823 |
| Qwen3.8 27B | 75·8 | 13·9 | 968 |
| Llama 3.3 70B | 70·6 | 19·1 | 255 |
| GLM-4.7-Flash | 69·1 | 73·3 | 3132 |

| | | | |
|---|---|---|---|
| Phi-4 14B | 64·1 | 3·1 | 187 |
| Gemma 3 27B | 63·2 | 15·3 | 309 |
| Llama 3.1 8B | 60·0 | 1·5 | 165 |
| Qwen2.5 7B | 58·0 | 1·2 | 171 |
| Gemma 3 12B | 57·9 | 6·1 | 327 |
| DeepSeek-R1 7B | 47·5 | 19·6 | 1727 |
| Mistral 7B | 46·1 | 0·8 | 127 |

##### Table S2: Truncated replies and recovery (all experiments)

| Model | Answers | Truncation retries | Graded on reasoning trace | Answered (%) |
|---|---|---|---|---|
| DeepSeek-R1 7B | 1250 | 138 | 52 | 96·6 |
| GLM-4.7-Flash | 1250 | 199 | 99 | 97·8 |
| Gemma 3 12B | 1850 | 0 | 0 | 99·5 |
| Gemma 3 27B | 3390 | 0 | 0 | 99·6 |
| Gemma 4 31B | 2040 | 108 | 45 | 98·1 |
| Llama 3.1 8B | 2790 | 0 | 0 | 99·4 |
| Llama 3.3 70B | 1890 | 0 | 0 | 99·6 |
| Mistral 7B | 1250 | 0 | 0 | 97·4 |
| Phi-4 14B | 1250 | 0 | 0 | 99·8 |
| Qwen2.5 7B | 1890 | 0 | 0 | 99·1 |
| Qwen2.5-VL 7B | 600 | 0 | 0 | 99·0 |
| Qwen3.8 27B | 2040 | 30 | 21 | 100·0 |

##### Table S3: Sensitivity analysis for recovered answers (experiment 1)

| Model | Flagged answers | Pooled accuracy as reported (%) | Flagged excluded (%) | Flagged counted incorrect (%) | Rank: reported / excluded / incorrect |
|---|---|---|---|---|---|
| Gemma 4 31B | 13 | 80·9 | 81·8 | 80·6 | 1 / 1 / 1 |
| Qwen3.8 27B | 1 | 75·8 | 75·9 | 75·8 | 2 / 3 / 2 |
| Llama 3.3 70B | 0 | 70·6 | 70·6 | 70·6 | 3 / 4 / 3 |
| GLM-4.7-Flash | 147 | 69·1 | 76·5 | 64·6 | 4 / 2 / 4 |
| Phi-4 14B | 0 | 64·1 | 64·1 | 64·1 | 5 / 5 / 5 |
| Gemma 3 27B | 0 | 63·2 | 63·2 | 63·2 | 6 / 6 / 6 |
| Llama 3.1 8B | 0 | 60·0 | 60·0 | 60·0 | 7 / 7 / 7 |
| Qwen2.5 7B | 0 | 58·0 | 58·0 | 58·0 | 8 / 8 / 8 |
| Gemma 3 12B | 0 | 57·9 | 57·9 | 57·9 | 9 / 9 / 9 |
| DeepSeek-R1 7B | 72 | 47·5 | 48·6 | 44·9 | 10 / 10 / 11 |
| Mistral 7B | 0 | 46·1 | 46·1 | 46·1 | 11 / 11 / 10 |

Flagged: answer extracted from the end of the reasoning trace because the reply was empty after the token limit, or reply obtained after a truncation retry (section 3). 950 medical questions per model.

##### Table S4: MedQA accuracy by protocol (experiment 2) and across repeats (experiment 3)

| Model | Protocol | Default (%) | Protocol (%) | Difference, points (95% CI) | Discordant questions | Holm-adjusted p |
|---|---|---|---|---|---|---|
| Gemma 3 27B | Letter-only | 74·0 | 72·0 | -2·0 (-6·8 to 2·8) | 13 | 1·00 |
| Gemma 3 27B | Chain-of-thought | 74·0 | 74·0 | 0·0 (-6·4 to 6·4) | 24 | 1·00 |

| | | | | | | |
|---|---|---|---|---|---|---|
| Gemma 3 27B | Three-shot | 74·0 | 67·3 | -6·7 (-13·7 to 0·4) | 30 | 0·79 |
| Llama 3.1 8B | Letter-only | 62·0 | 57·3 | -4·7 (-13·6 to 4·4) | 49 | 1·00 |
| Llama 3.1 8B | Chain-of-thought | 62·0 | 71·3 | 9·3 (1·0 to 17·5) | 42 | 0·39 |
| Llama 3.1 8B | Three-shot | 62·0 | 58·0 | -4·0 (-13·2 to 5·4) | 52 | 1·00 |
| Llama 3.3 70B | Letter-only | 84·0 | 83·3 | -0·7 (-6·3 to 4·9) | 17 | 1·00 |
| Llama 3.3 70B | Chain-of-thought | 84·0 | 87·3 | 3·3 (-1·5 to 8·4) | 13 | 1·00 |
| Llama 3.3 70B | Three-shot | 84·0 | 85·3 | 1·3 (-3·7 to 6·5) | 14 | 1·00 |
| Qwen2.5 7B | Letter-only | 68·0 | 60·7 | -7·3 (-13·1 to -1·5) | 21 | 0·27 |
| Qwen2.5 7B | Chain-of-thought | 68·0 | 56·7 | -11·3 (-18·4 to -4·0) | 33 | 0·05 |
| Qwen2.5 7B | Three-shot | 68·0 | 57·3 | -10·7 (-17·7 to -3·4) | 32 | 0·08 |

Same 150 MedQA questions; Newcombe hybrid score CI for the paired difference; exact McNemar test, Holm correction across the 12 comparisons.

| Model | Temperature | Mean (%) | SD | Min | Max | Identical outcome (%) |
|---|---|---|---|---|---|---|
| Gemma 3 27B | 0·0 | 74·0 | 0·00 | 74·0 | 74·0 | 100·0 |
| Gemma 3 27B | 0·7 | 74·9 | 0·38 | 74·7 | 75·3 | 98·7 |
| Llama 3.1 8B | 0·0 | 62·0 | 0·00 | 62·0 | 62·0 | 100·0 |
| Llama 3.1 8B | 0·7 | 54·9 | 4·07 | 51·3 | 59·3 | 59·3 |

#### Table S5: Image ablation (experiment 4)

| Model | Benchmark | With image (%) | Without image (%) | Gain, points (95% CI) | Correct only with image | Correct only without | Holm-adjusted p |
|---|---|---|---|---|---|---|---|
| Gemma 3 12B | MedXpertQA MM | 20·7 | 24·7 | -4·0 (-12·2 to 4·3) | 17 | 23 | 1·00 |
| Gemma 3 12B | MMMU medical | 50·0 | 40·7 | 9·3 (0·4 to 18·0) | 31 | 17 | 0·48 |
| Gemma 3 27B | MedXpertQA MM | 32·0 | 27·3 | 4·7 (-3·5 to 12·7) | 23 | 16 | 1·00 |
| Gemma 3 27B | MMMU medical | 53·3 | 44·0 | 9·3 (0·4 to 18·0) | 31 | 17 | 0·48 |
| Gemma 4 31B | MedXpertQA MM | 55·3 | 49·3 | 6·0 (-1·9 to 13·7) | 23 | 14 | 0·94 |
| Gemma 4 31B | MMMU medical | 66·7 | 48·7 | 18·0 (10·0 to 25·6) | 34 | 7 | 0·0002 |
| Qwen2.5-VL 7B | MedXpertQA MM | 18·0 | 19·3 | -1·3 (-8·1 to 5·4) | 12 | 14 | 1·00 |
| Qwen2.5-VL 7B | MMMU medical | 49·3 | 42·7 | 6·7 (-0·9 to 14·0) | 22 | 12 | 0·73 |
| Qwen3.8 27B | MedXpertQA MM | 56·0 | 50·0 | 6·0 (-3·4 to 15·3) | 31 | 22 | 1·00 |
| Qwen3.8 27B | MMMU medical | 75·3 | 56·0 | 19·3 (11·7 to 26·7) | 34 | 5 | <0·0001 |

Same 150 questions per benchmark with and without images; gain = accuracy with image minus accuracy without image (positive values mean the image helped). Newcombe hybrid score CI for the paired difference; exact McNemar test, Holm correction across the ten comparisons. Gemma 3 12B and Gemma 3 27B have the same discordant counts on MMMU (31 and 17) on largely different questions (8 and 5 shared), so their CIs and p values coincide.

#### Table S6: Agentic computational biology (experiment 5)

| Model | BioCompute closed-book (%) | BioCompute with tools (%) | Gain, points (95% CI) | Holm-adjusted p | BioAgent (%) |
|---|---|---|---|---|---|
| Gemma 3 27B | 6·7 | 69·3 | 62·7 (48·6 to 72·9) | <0·0001 | 7·5 |
| Gemma 4 31B | 38·7 | 96·0 | 57·3 (44·1 to 67·8) | <0·0001 | 100·0 |
| Llama 3.1 8B | 4·0 | 57·3 | 53·3 (38·6 to 64·8) | <0·0001 | 2·5 |
| Llama 3.3 70B | 17·3 | 53·3 | 36·0 (20·5 to 49·2) | <0·0001 | 5·0 |
| Qwen2.5 7B | 5·3 | 41·3 | 36·0 (22·6 to 48·0) | <0·0001 | 2·5 |

| | | | | | |
|---|---|---|---|---|---|
| Qwen3.8 27B | 56·0 | 98·7 | 42·7 (31·2 to 53·8) | <0·0001 | 97·5 |

| Model | Benchmark | Mean turns | Mean tool calls | Mean tool errors | Unanswered | Items |
|---|---|---|---|---|---|---|
| Gemma 3 27B | bioagent | 5·1 | 2·6 | 0·4 | 4 | 40 |
| Gemma 3 27B | biocompute_agent | 2·1 | 0·9 | 0·0 | 0 | 75 |
| Gemma 4 31B | bioagent | 6·7 | 5·7 | 0·0 | 0 | 40 |
| Gemma 4 31B | biocompute_agent | 2·7 | 1·6 | 0·0 | 0 | 75 |
| Llama 3.1 8B | bioagent | 2·5 | 12·0 | 0·0 | 3 | 40 |
| Llama 3.1 8B | biocompute_agent | 2·7 | 3·4 | 0·1 | 0 | 75 |
| Llama 3.3 70B | bioagent | 2·0 | 1·1 | 0·0 | 2 | 40 |
| Llama 3.3 70B | biocompute_agent | 1·7 | 0·8 | 0·0 | 0 | 75 |
| Qwen2.5 7B | bioagent | 7·8 | 47·6 | 12·9 | 7 | 40 |
| Qwen2.5 7B | biocompute_agent | 1·9 | 13·8 | 0·1 | 4 | 75 |
| Qwen3.8 27B | bioagent | 4·7 | 4·1 | 0·0 | 0 | 40 |
| Qwen3.8 27B | biocompute_agent | 2·4 | 1·4 | 0·0 | 0 | 75 |

| Task family | Closed-book (%) | With tools (%) |
|---|---|---|
| ts tv ratio | 36·1 | 38·9 |
| cigar span | 47·9 | 45·8 |
| motif positions | 0·0 | 50·0 |
| snv effect | 59·5 | 52·4 |
| mean phred | 25·0 | 55·6 |
| frequent kmer | 8·3 | 66·7 |
| protein mw | 12·5 | 75·0 |
| primer tm | 37·5 | 79·2 |
| global alignment | 8·3 | 80·0 |
| restriction count | 13·3 | 83·3 |
| reverse complement | 0·0 | 91·7 |
| translate cds | 0·0 | 91·7 |
| gc content | 7·1 | 95·2 |
| longest orf | 0·0 | 95·8 |

#### Table S7: Python interpreter versus sequence tools (experiment 5)

| Model | Benchmark | Python calls | Sequence-tool calls | Other calls | Items using Python | Python share of calls (%) |
|---|---|---|---|---|---|---|
| Gemma 3 27B | bioagent | 12 | 48 | 45 | 6/40 | 11·4 |
| Gemma 3 27B | biocompute_agent | 10 | 61 | 0 | 9/75 | 14·1 |
| Gemma 4 31B | bioagent | 143 | 10 | 76 | 40/40 | 62·5 |
| Gemma 4 31B | biocompute_agent | 60 | 50 | 8 | 34/75 | 50·9 |
| Llama 3.1 8B | bioagent | 478 | 0 | 3 | 37/40 | 99·4 |
| Llama 3.1 8B | biocompute_agent | 15 | 228 | 15 | 14/75 | 5·8 |
| Llama 3.3 70B | bioagent | 34 | 5 | 3 | 30/40 | 81·0 |
| Llama 3.3 70B | biocompute_agent | 12 | 32 | 12 | 11/75 | 21·4 |
| Qwen2.5 7B | bioagent | 5 | 1342 | 556 | 2/40 | 0·3 |
| Qwen2.5 7B | biocompute_agent | 2 | 1018 | 14 | 1/75 | 0·2 |
| Qwen3.8 27B | bioagent | 70 | 18 | 76 | 40/40 | 42·7 |

| | | | | | | |
|---|---|---|---|---|---|---|
| Qwen3.8 27B | biocompute_agent | 40 | 44 | 18 | 29/75 | 39·2 |

| Benchmark | Tools used in the item | Items | Accuracy (%) |
|---|---|---|---|
| bioagent | both | 17 | 88·2 |
| bioagent | neither | 60 | 3·3 |
| bioagent | python only | 138 | 49·3 |
| bioagent | sequence tools only | 25 | 4·0 |
| biocompute_agent | both | 28 | 96·4 |
| biocompute_agent | neither | 121 | 30·6 |
| biocompute_agent | python only | 70 | 80·0 |
| biocompute_agent | sequence tools only | 231 | 83·1 |

Counts from the stored trajectories. Sequence tools: reverse complement, translation, GC content, ORF search, restriction sites, alignment, protein mass and primer Tm. Other: calculator, file listing and reading. Accuracy by tool use is descriptive: models chose which tools to use.

#### 9. Reproducing the study

The experiment plan (`paper_experiments/plan.py`) lists models, benchmarks, sample sizes and the seed. With the platform installed and the benchmarks loaded (see README):

```
python manage.py runserver 0.0.0.0:8020
python manage.py bm_provider add paper-ollama "Paper experiments (Ollama)" --type ollama --url http://<host>:11434
python paper_experiments/run_plan.py                  # submits every run through the REST API
python manage.py bm_regrade --tag paper2026           # scores all stored replies with the final rules
python manage.py bm_manifest --tag paper2026 --out paper_experiments/data/manifest
python paper_experiments/export.py                    # runs, per-question results and raw replies
python paper_experiments/validate_generated.py        # independent ground truth
python paper_experiments/extraction_audit.py final --legacy <grades from commit 9d03117>
jupyter nbconvert --to notebook --execute --inplace paper_experiments/notebooks/*.ipynb
```

Questions are drawn with seed 2026, so every installation evaluates the same items; the fingerprints in section 2 confirm it.
